# Rotation-based schedules in elementary schools to prevent COVID-19 spread: A simulation study

**DOI:** 10.1101/2021.06.28.21259628

**Authors:** Cyril Brom, Tomáš Diviák, Jakub Drbohlav, Václav Korbel, René Levínský, Roman Neruda, Gabriela Suchopárová, Josef Šlerka, Martin Šmíd, Jan Trnka, Petra Vidnerová

## Abstract

**Background:** Rotations of schoolchildren on a weekly basis is one of the nonpharmaceutical interventions often considered in the covid-19 pandemic. This study aims to investigate the impact of different types of rotations in various testing contexts.

**Methods:** We built an agent-based model of interactions among pupils and teachers based on an online survey in an elementary school in Prague, Czechia. This model contains 624 schoolchildren and 55 teachers (679 nodes) and about 27 thousands social contacts (edges) in 10 layers. The layers reflect different types of contacts (in classroom, cafeteria etc.) as described in the survey. On this multi-graph structure we run a modified SEIR model of the covid-19 dynamics. The parameters of the model are calibrated on data from the outbreak in the Czech Republic in the period March to June 2020.

**Findings:** There are three main findings in our paper.

1. Weekly rotations of in-class and distance learning reduce the spread of covid-19 by 75–81% and thus represent an effective preventative measure in school setting.
2. Regular antigen testing twice a week, or weekly PCR testing, significantly reduces infections even when using tests with a lower sensitivity: tests with a 40% sensitivity reduce infections by more than 50 percent.
3. The density of revealed contact graphs for older pupils is 1.5 times higher than the younger pupils graph, the teachers network is yet an order of magnitude denser. Consequently, the infection transmission between teachers is highly overproportional in our school. Moreover, teachers act as bridges connecting clusters of classes, especially in the secondary grade where they are responsible for 14–18% of infections, in comparison to 8–11% in primary grade.

**Interpretation:** Weekly rotations with regular testing are a highly effective non-pharmaceutical intervention for the prevention of covid-19 spread in schools and a way to keep schools open during an epidemic or to reopen them as the epidemiological situation improves.

## 1 Introduction

Interpersonal contacts are the main channel of transmission of the SARS-CoV-2 coronavirus. The transmission may occur in many locations of human activity such as households, shops, or workplaces. Schools in general are one of the major environments where the virus may spread and infect large numbers of individuals both within and outside of a particular school (Gold, 2021; Stein-Zamir et al., 2020; Torres et al., 2020); see also Forbes et al. (2021); Ismail et al. (2021); Lessler et al. (2021); SAGE (2020); Brauner et al. (2021); Haug et al. (2020), but also Zhu et al. (2020); Zimmerman et al. (2021).

School closures were therefore among the first large-scale epidemic control measures during the covid-19 pandemic. Since the spring of 2020, schools all over the world have been either closed for in-person education or operating under some restrictive measures. For instance, in Czechia, where we based our study, schools closed in mid-March 2020, briefly reopened in September 2020 only to close down again in October 2020 due to the rapid increase in the number of newly detected cases. With some exceptions affecting only small numbers of students and over short periods of time, Czech schools remained closed until the middle of April 2021.

The reopening of schools amidst an ongoing pandemic poses a distinct challenge. On the one hand, school reopening may expose pupils, their families, teachers, and other school staff to the risk of contracting the infection and subsequently spreading it further (Forbes et al., 2021; Lessler et al., 2021; SAGE, 2020); see also Brauner et al. (2021); Haug et al. (2020), but also Zhu et al. (2020). Even though covid-19 is unlikely to cause severe disease in children, estimates of the prevalence of long covid symptoms based, e.g. in the UK (ONS, 2021) suggest that 13% of children aged 2–10 years and 15% of those aged 12–16 years have at least one persistent symptom 5 weeks after testing positive (Gurdasani et al., 2021).

On the other hand, there is evidence that a long-term lack of contacts with peers may negatively affect children’s psychological development and well-being (Bignardi et al., 2020; ECDC, 2020; Di Pietro et al., 2020; Ravens-Sieberer et al., 2021), but see also Hafstad et al. (2021); as well as their educational outcomes (Di Pietro et al., 2020; Engzell et al., 2020; Maldonado and De Witte, 2020), with home-schooling and online learning disproportionately negatively affecting pupils and students from disadvantaged social backgrounds (Di Pietro et al., 2020; ECDC, 2020).

Despite the importance of re-opening schools, there is a lack of empirical evidence on how to do so safely, not least because it is hardly conceivable to conduct experiments in schools during the covid-19 pandemic and it is very difficult to acquire sufficiently granular data from epidemiological records.

One way to gain insight from imperfect empirical data is to leverage a combination of agent-based simulations with epidemiological models on empirical contact networks. The structure of contact networks relevant to the spread of a particular pathogen affects the extent and the dynamics of its spread in the given population (Danon et al., 2011; Luke and Harris, 2007). This has also been recognized in previous research focusing on the structure of contact networks within schools and their effect on transmission of respiratory infections such as influenza (McGee et al., 2021; Gemmetto et al., 2014; Stehlé et al., 2011).

In this study we contribute to the existing research by utilizing empirical data to build a network of interpersonal contacts within an elementary school and use it in agent-based simulations of the SARS-CoV-2 transmission. To our knowledge this is the first such study. Other modelling studies have been published (Phillips et al., 2021; McGee et al., 2021; Panovska-Griffiths et al., 2020) but none uses empirical data on interpersonal contacts among pupils. We use simulations to assess the effect of a range of schedules and measures on the extent and dynamics of the epidemic in a school setting. We investigate the effect of rotation (i.e. alternating groups of pupils that are physically present in school) and testing. We thus obtain realistic results that would otherwise be impossible to obtain due to ethical and practical considerations. The results of our model have already been leveraged by the Czech government to construct a set of measures to reopen schools in April 2021.

## 2 Methods

### 2.1 Data

To accurately capture the network of contacts between all individuals, we conducted a questionnaire survey in a selected elementary school in Czechia, which educates pupils in grades 1-9 (1-5 primary school, 6-9 lower secondary school). In this survey, we asked both pupils and teachers with whom, where, and how often they interacted. The survey was conducted online after the schools were closed for the second time in Czechia on November 2, 2020 (school closure started on October 14, 2020). For the full questionnaire, further details about data collection, and descriptive analysis, see the Appendix.

Data from the survey described a multilevel multiplex network with two types of nodes: pupils and teachers. Six types of contacts among pupils relevant to SARS-CoV-2 transmission were studied in the survey: sitting next to each other at a desk; contacts during breaks between classes; contacts during lunch break; contacts in after-school care facilities; contacts during school-based voluntary activities; and contacts outside the school. Additional three types of relevant contacts among teachers were examined in the survey: contacts in a shared office; contacts within the school but outside the office; and contacts outside the school. The intensity of all these types of contacts was measured on a four-point scale ranging from 0 (no contact) to 4 (multiple times a day). The last type of contacts are contacts that connect the teachers and the pupils during classes.

### 2.2 Model

An agent-based network model consists of three components: a realistic synthetic population, its social contact network, and an epidemiological SEIR model (Eubank et al., 2004). The population and the contacts are directly built from the collected data as described above. The epidemiological model consists of our extension of the SEIR model applied to an agent-based model of a Czech municipality described in (Berec et al., 2021). Basic disease-related parameters were taken from this model calibrated to the pandemic situation in Czechia in March-June 2020. The probability of infection in our model depends on three factors: the disease infection rate, contact intensity, and the level of individual protection. The infection rate is taken directly from our calibrated model as 0.32 for symptomatic individuals and 0.16 for asymptomatic ones. Contact intensities for various school environments are also taken from our previous research. They take into account contact duration and the distance and environment at which the contact takes place. The level of covid-19 epidemic in the society is reflected in our model by random daily infection imports. Details of the model and simulation parameters are presented in the Appendix.

### 2.3 Simulated measures

We model the impact of two measures, weekly rotation of in-class and distance learning, regular testing of pupils and teachers, and the combination thereof. We look at the spread of covid-19 at grades 1-5 (primary education, typically children aged 6-11 years) and grades 6-10 (lower secondary education, typically children aged 11-15 years).

For weekly rotation, we consider alterations of entire classes without any reduction in the number of pupils in the class, i.e. all pupils in a given class attend in-class learning in week 1, followed by distance learning in week 2. A scenario where all classes are divided into halves and these halves alternate on a weekly basis is also considered to estimate the impact of in-class contacts.

For different testing regimes, we consider weekly PCR tests with a conservative 80% sensitivity and antigen tests with sensitivities of 10%, 20% and 40% performed once or twice a week. In both regimes we assume that positive pupils won’t infect other pupils after taking the test and that they don’t continue to attend school after testing positive. We don’t assume any other quarantine or isolation measures for the rest of the class.

**Panel 1:** Simulation scenarios overview. Basic scenarios serve as baseline for further experiments and they also estimate the relative role of pupils in primary and lower secondary grades and the role of teachers. Rotation scenarios compare different versions of class-based weekly rotation and rotation of a half of a class. Testing scenarios consider various testing efficiency and performance parameters.

|  |  |
| --- | --- |
| **Basic scenarios** |  |
| Baseline | everything open except after-school clubs |
| Closed | the whole school is closed |
| G1–5 full, G6–9 closed | only grades 1–5 are attending |
| G1–5 closed, G6–9 full | only grades 6–9 are attending |
| no teachers | theoretical scenario where teachers never spread infection |
| **Rotation scenarios** |  |
| Rotations | all groups are attending school in a weekly rotation manner |
| Half rotations | each group is divided in half, these halves are attending school in weekly rotation manner, i.e. each week, half of each group at home, the other half at school |
| G1-5 rotate, G6-9 closed | grades 1-5 are attending school in rotations, grades 6–9 are at home |
| G1–5 full, G6–9 rotate | grades 1–5 are attending school daily, grades 6–9 in a rotation manner |
| **Testing scenarios** |  |
| PCR test once | students are tested on Sundays by PCR test (0.8 sensitivity) |
| Antigen test twice | students are tested on Mondays and Thursdays by antigen test (0.1/0.2/0.4 sensitivity) |
| Antigen test once | students are tested on Mondays by antigen test (0.1/0.2/0.4 sensitivity) |

## 3 Results

### 3.1 Scenarios

All scenarios used in our simulations are summarized in Panel 1 divided into three categories. The baseline scenarios model situations where the school is open (except for after-school clubs), completely closed, or where only primary grades or only secondary grades pupils attend. The last two variants were also considered in an artificial scenario without teachers present at school, in order to estimate the contribution of teachers to the spread of the infection (this case corresponds to a hypothetical scenario of fully immune, noninfectious teachers).

Rotation scenarios comprise weekly rotation of primary grades pupils (secondary grades are closed), secondary grades rotation (primary grades without restrictions), weekly rotation of all classes, and weekly rotation of halves of classes. The testing scenarios comprise highly sensitive PCR testing performed on Sundays or antigen testing using tests of various sensitivity performed either once or twice a week (on Mondays, or Monday and Thursday mornings, respectively).

All simulations were run at four different strengths of the epidemic characterized by daily infected imports corresponding to 146 per 100 000 people (what we refer to as severe epidemic), 73 (50% of the severe epidemic) 36 (25%), and 15 (10%). All simulation results can be found in the Appendix.

We used the simulations to compare three effects: the effect of rotation only, the effect of testing only, and the combined effect of rotation and testing.

### 3.2 Rotations

Weekly rotation has a major impact on curbing the spread of the infection within the school (Figure 1 and Table 1). When all groups of pupils take part in rotation schedules the infection rate drops to 19–25% compared to a fully open school. When the school opens only for primary grades, alternatively introducing also rotation, the infection rate drops to 51–54% or to mere 12–15%, respectively, compared to the infection rate of a fully open school. Rotation of halves of all classes is comparably efficient (13–15%).

**Table 1:** Rotation scenarios. Percentage of infected cases compared to the baseline six weeks after the start of the policy. Mean values of 1000 runs are reported. (Baseline, i.e. no restrictions, corresponds to 100%, closed schools corresponds to 0%).

|  | import 0.1 | import 0.25 | import 0.5 | import 1.0 |
| --- | --- | --- | --- | --- |
| G1–5 full, G6–9 closed | 50.67 | 54.67 | 52.98 | 54.38 |
| G1–5 full, G6–9 rotate | 60.19 | 63.87 | 63.92 | 64.46 |
| rotations | 18.93 | 21.94 | 22.69 | 24.79 |
| half rotations | 13.18 | 12.57 | 14.02 | 15.42 |
| G1–5 rotate, G6–9 closed | 12.17 | 12.47 | 13.83 | 15.32 |

**Figure 1:**
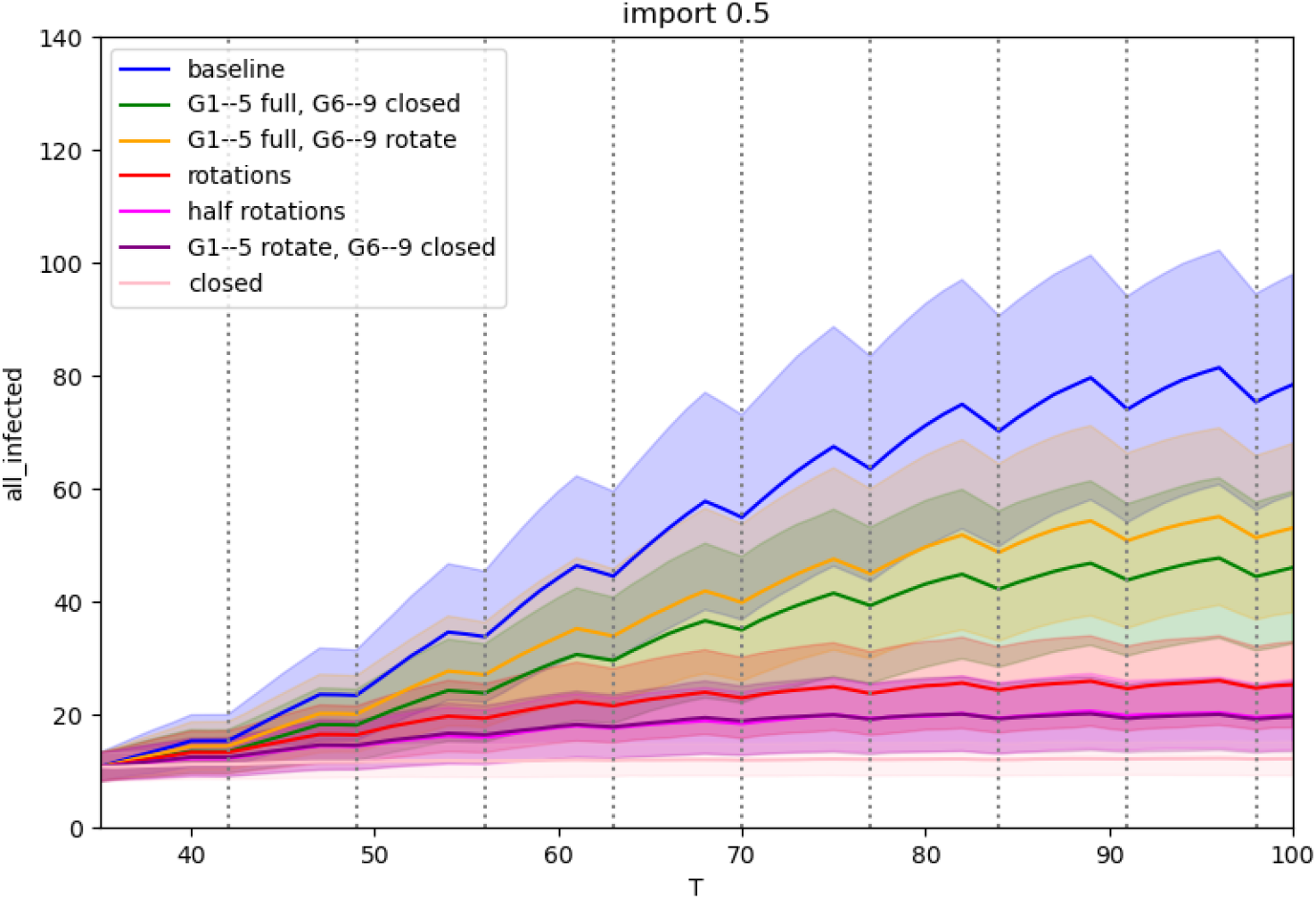
Rotation scenarios for medium epidemics: Comparison of the number of infected active cases during the run. The x-axis represents days of simulation, the y-axis represents the number of infections at school, mean values of 1000 runs with standard deviation are plotted for each simulation.

### 3.3 Testing

The impact of testing depends heavily on the sensitivity of the testing procedure (Figure 2 and Table 2). Highly sensitive PCR tests are able to reduce infection rates down to 38–45%, while low sensitivity antigen tests have a much lower impact even when performed twice a week (from 43% of infections compared to no intervention for 0.4 sensitivity tests and a mild epidemic to 85% of infections for 0.1 sensitivity tests and a severe epidemic). For comparison antigen tests performed once a week are also included in the experiments. Unsurprisingly, their effect is even lower (61% of infections in contrast to 43%, and 92% in contrast to 85% with corresponding epidemic level and sensitivities).

**Table 2:**
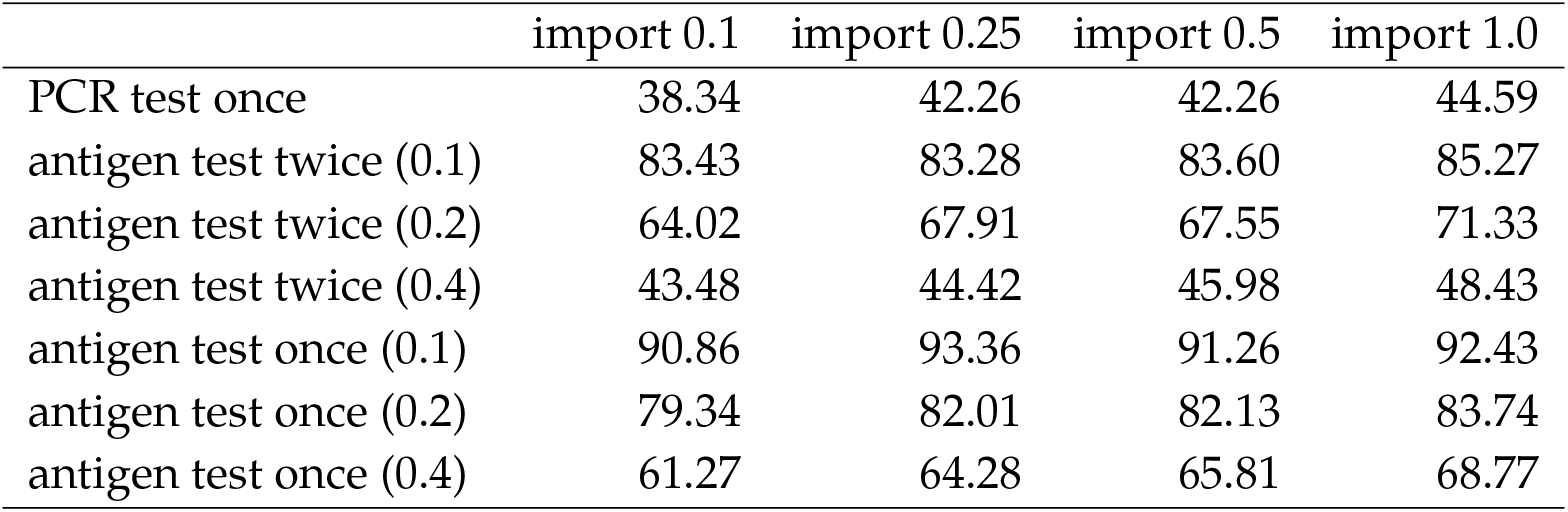
Testing scenarios. Number of infected cases compared to baseline six weeks after the start of the policy expressed as a percentage. The numbers in brackets denote the sensitivity of the test. (Baseline, i.e. no restrictions, corresponds to 100%, closed schools correspond to 0%).

**Figure 2:**
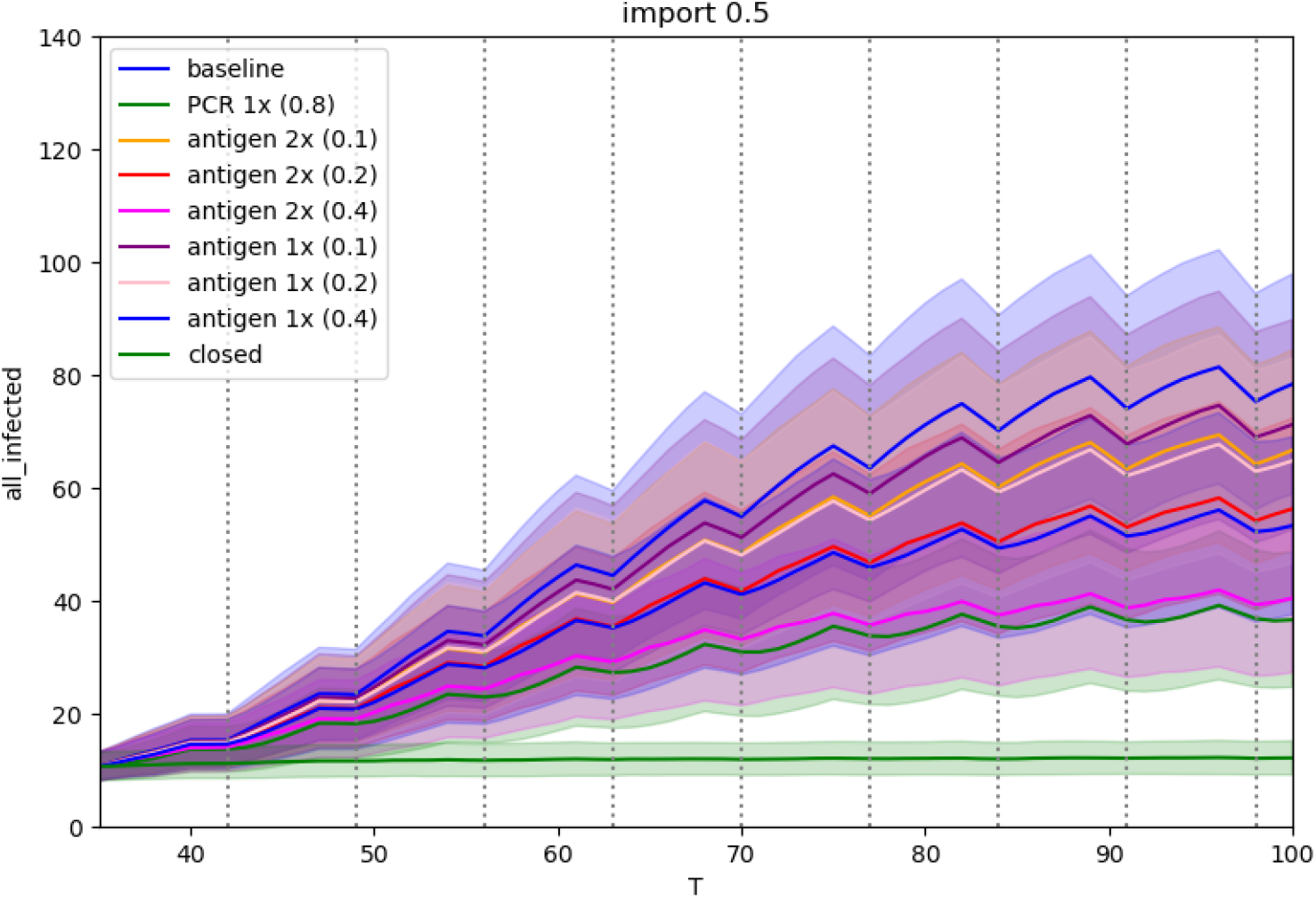
Testing scenarios for a medium epidemic: Comparison of the number of infected active cases during the run. The x-axis represents days of simulation, the y-axis represents number of infections at school, mean values of 1000 runs with standard deviation are plotted for each simulation.

### 3.4 Combined measures

The already strong effect of rotation is further amplified by adding testing (Figure 3 and Table 3). The combination of rotations and sensitive PCR tests drops infection numbers to the range of mere 13–15% compared to no restrictions, and even low sensitivity antigen tests reduce infections to 15–23% when performed twice a week, and 17–24% when performed once a week.

**Table 3:** Rotation + testing scenarios. Number of infected cases compared to baseline six weeks after the start of the policy expressed as a percentage. Mean values of 1000 runs are reported. (Baseline, i.e. no restrictions, corresponds to 100%, closed schools correspond to 0%).

|  | import 0.1 | import 0.25 | import 0.5 | import 1.0 |
| --- | --- | --- | --- | --- |
| rotation + PCR test once | 12.77 | 13.12 | 12.95 | 14.76 |
| rotation + antigen test twice (0.1) | 18.46 | 19.45 | 19.76 | 22.50 |
| rotation + antigen test twice (0.2) | 16.65 | 16.96 | 17.87 | 20.51 |
| rotation + antigen test twice (0.4) | 14.76 | 14.11 | 14.25 | 16.28 |
| rotation + antigen test once (0.1) | 19.51 | 20.24 | 20.61 | 23.63 |
| rotation + antigen test once (0.2) | 18.70 | 19.51 | 19.80 | 22.23 |
| rotation + antigen test once (0.4) | 16.92 | 16.38 | 16.94 | 19.49 |

**Figure 3:**
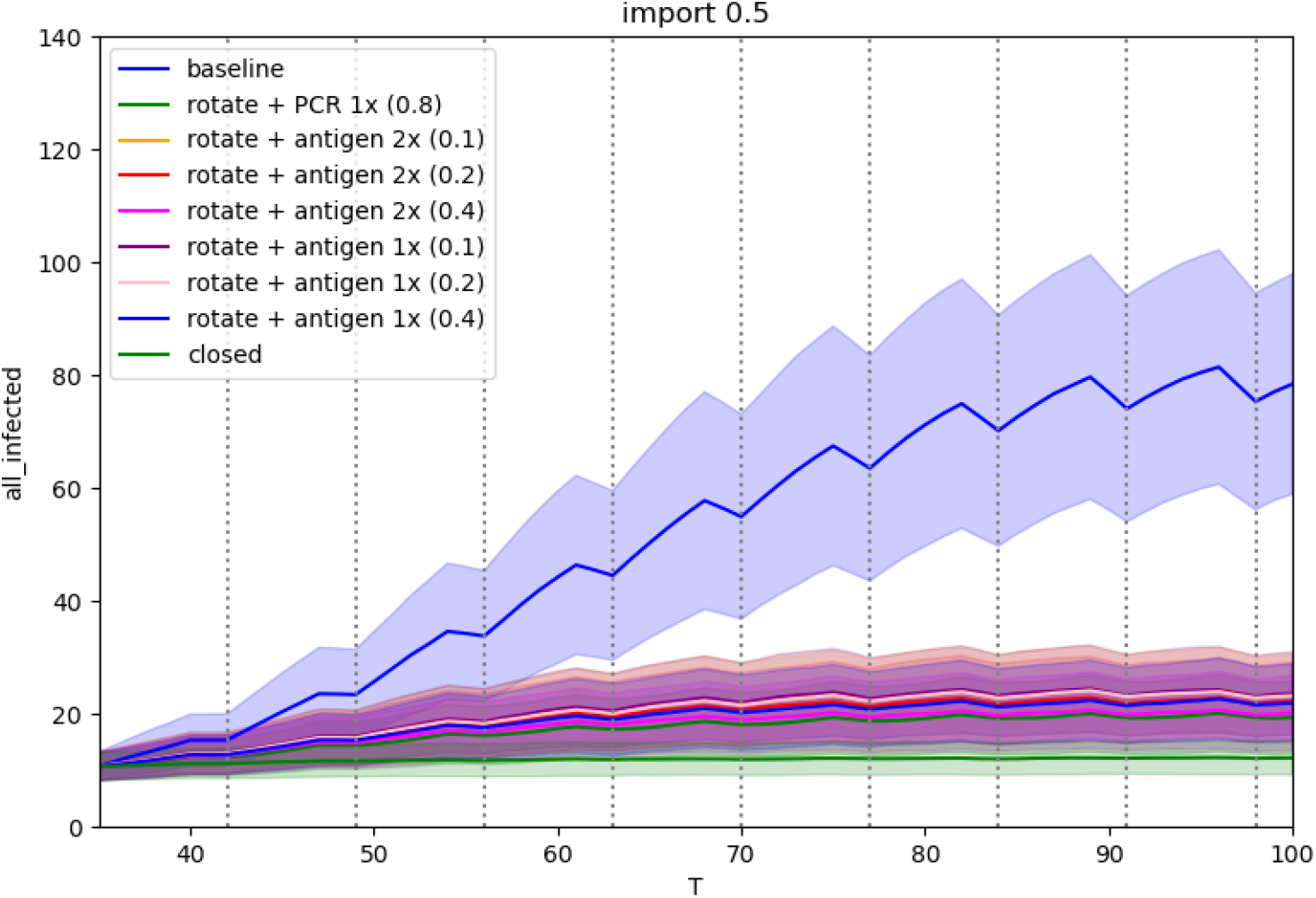
Rotations and testing scenarios for a medium epidemic: Comparison of the number of infected active cases during the run. The x-axis represents days of simulation, the y-axis represents number of infections at school, mean values of 1000 runs with standard deviation are plotted for each simulation.

### 3.5 Role of teachers in spreading the infection

Statistics of the sources of infection in simulations show that contacts between teachers represent almost 6% of school infections. However, teachers represent only 8% of individuals in our school, i.e. they would be responsible only for 0.64% in a naïve symmetric graph model. The fact that we find the infection transmission between teachers almost ten times higher compared to a symmetric graph is a direct consequence of the revealed structure of social interactions in our school. The study of relative densities of contact graphs reveals that the density of older pupils subgraph is 1.5 times higher than the younger pupils subgraph. The teachers subgraph has yet an order of magnitude higher density (cf. Appendix).

The scenarios in Table 4 compare a completely open school (taken as 100% infection rate) with the attendance of grades 1–5 only, resulting in infection rates between 51–54%, and with the attendance of grades 6–9 only, resulting in 44–46% infection rate. Then an artificial scenario assumes the same attendance with fully immune, noninfectious teachers. We can see that the ‘absence’ of teachers reduces the infections by 8–11% for grades 1–5 and by 14–18% for grades 6–9. These results show a bigger role of teachers for the viral spread in grades 6–9 due to their teaching in multiple classes in these grades.

**Table 4:** The role of teachers. Numbers of infected cases compared to the baseline six weeks after the start of the policy expressed as a percentage. Mean values of 1000 runs are reported. (Baseline, i.e. no restrictions, corresponds to 100%, closed schools correspond to 0%).

|  | import 0.1 | import 0.25 | import 0.5 | import 1.0 |
| --- | --- | --- | --- | --- |
| G1–5 full, G6–9 closed | 50.67 | 54.66 | 52.98 | 54.38 |
| G1–5 full, G6–9 closed, no teachers | 42.29 | 44.12 | 44.13 | 44.61 |
| difference for G1–5 | 8.38 | 10.54 | 8.85 | 9.77 |
| G1–5 closed, G6–9 full | 45.35 | 44.24 | 45.91 | 44.50 |
| G1–5 closed, G6–9 full, no teachers | 27.22 | 28.47 | 29.79 | 30.24 |
| difference for G6–9 | 18.13 | 15.77 | 15.98 | 14.26 |

## 4 Discussion

The results of our model indicate that weekly rotation of in-class and distance learning can be very effective in curbing the spread of covid-19 infection in elementary schools (by as much as 75–81%). As the modelled rotation does not assume any changes to the composition of the class, it brings about only a limited disturbance to a normal school operation. The effectiveness of rotation is insensitive to individual compliance within schools, often a key factor in the effectiveness of other protective measures focused on individuals, such as wearing protective masks or heightened hygienic measures.

Our results also indicate a strong effect of regular testing of pupils in schools, even with lower sensitivity tests, provided a negative test is a requirement to participate in in-class education and the tests are performed twice a week several days apart in the case of lower sensitivity tests. Antigen tests of a 40% sensitivity performed twice a week reduce the spread of covid-19 by 52–57%. For comparison, PCR tests (with an 80% sensitivity) performed once a week reduce the spread similarly by 55–62%, while antigen tests (40% sensitivity) performed once a week result in about 31–39% reduction. The combined effect of rotation and testing results in a strong 84–86% reduction of infections for antigen tests with a 40% sensitivity performed twice a week. Our results also show that teachers play a key role in the spread of covid-19 in schools, especially in the secondary school grades. The contribution to reduction of in infection rates by excluding teachers from school represents is 8–11% for primary grades, but 14–18% for secondary grades. This is roughly in line with observation studies indicating that teachers tend to be overrepresented among secondary cases in schools compared to students (see, e.g., Gold, 2021; Ismail et al., 2021; Schoeps et al., 2021), though there is some heterogeneity in the literature, with the reported raw ratio of teachers:students being approx. 1:2 (Schoeps et al., 2021), but also approx. 1:5 (Gold, 2021).

An additional analysis of contact networks of children in grades 1–5 compared to children in grades 6–9 indicates that the older children have about 1.5 times more contacts among themselves and also more contacts with teachers (cf. Appendix, Table 3A). While contacts from the same class are comparable for grades 1–5 and 6–9, the contacts with other classes are two times more common for older children. This is generally in line with the observation that school infection clusters among older children tend to be more frequent than among younger children (ECDC, 2020), see also (SAGE, 2020).

As any other model or simulation, our model is built on certain assumptions. One such assumption relates to the behavior of a person tested with a negative test result. We assume that a false negative test result does not alter behavior of the tested person in a significant way compared to behavior with no test at all. Further research on behavioral changes in children and teachers as a result of negative covid-19 test should be conducted to improve our findings. In a similar vein, we assume that individuals in our simulations are entirely compliant with the anti-epidemic measures imposed upon them (such as isolation following a positive test). This assumption may not necessarily hold in all situations, for example after a prolonged lockdown and restrictions. In these situations, the population may experience compliance fatigue, which may lower its willingness to comply with the mitigation measures. If the compliance is low, simulation models including the one we present here are likely to yield less valid results unless they specifically account for these negative effects. Further research may focus on how sensitive our results are to a lower compliance.

Other limitations of our study relate to the virological properties of the virus. Specifically, we are not considering different variants of SARS-CoV-2 and their effect in the epidemiological component of our SEIR model. The model was calibrated based on the Czech epidemiological situation from March to June 2020. However, different epidemic levels in our simulations, spanning an order of magnitude in infected imports, indicate the robustness of our results. Additionally, the model may be extended by calibrating its parameters to different variants of the SARS-CoV-2 virus or even to different viruses altogether provided that the relevant transmission pathways are interpersonal contact networks such as with other respiratory diseases.

Our results indicate the roles of specific components of elementary school operation in the spread of covid-19 and the effectiveness of selected measures and school regimes on the spread of covid-19 within schools. They do not pertain in any way to the role of schools themselves in the spread of covid-19 in the entire population.

To our knowledge, this is the first agent-based model assessing the effectiveness of individual measures on covid-19 spread in a school setting based on real contact networks derived from empirical data. Our contact network data align with previous research based on both simulated data (McGee et al., 2021) or contact networks obtained using wearable sensors (Gemmetto et al., 2014; Stehlé et al., 2011) in showing densely interconnected contact networks with strong within-class mixing among pupils. Likewise, the results obtained from our simulations point at similarities with other studies. Those include the disproportionate effect of only a subset of pupils not attending the school on decreasing the infection rates (Gemmetto et al., 2014) and in general, the effects of rotation on curbing the viral transmission (McGee et al., 2021; Panovska-Griffiths et al., 2020; Head et al., 2021), and the effect of regular testing even with low sensitivity tests on reducing the infection rates in a study with a similar design on artificial data (McGee et al., 2021) as well as in a population-level study (Panovska-Griffiths et al., 2020).

Our study thus presents the first assessment of selected school regimes and measures based on empirical data. As such it can serve not only to expand our knowledge of covid-19 transmission but it can also be used by public health authorities and policy makers for informed decisions on safe school operations during the covid-19 epidemic. The Czech Government has implemented weekly rotations for primary schools on the 12th of April 2021 for grades 1-5 and on the 3rd of May for grades 6-9 in most regions. The rotations are is accompanied by testing (twice a week for an antigen test and once a week for PCR test). While our results may be used for formulation drafting and implementation of policies in schools, policy-makers and other actors involved in decision making should always consider the broader epidemiological context and situation.

## Data Availability

Data and software used in the paper is available for download.

https://github.com/epicity-cz/model-m/releases/tag/v0.2-schools

## Appendix

### A1 Introduction

In this Appendix we have collected supplementary material – data, detailed results, and questionnaires – used in our study. The structure of the Appendix is as follows, in section A2 we describe the school and the contact networks created from the survey. Complete results in the form of tables and graphs with sensitivity analysis are gathered in section A3. Finally, sections A4 and A5 contain transcripts of electronic questionnaires used to collect the contact data from students and teachers, respectively.

### A2 Data

#### Collection and preprocessing

The school where we collected data is located in a suburban area of a large Czech city. The school has nine grades of primary and lower secondary education (children age 6 to 15). In total, 624 students and 55 teachers were included in the data collection. The school is representative of suburban schools in the Czech republic. In particular, the school has one main building where both the primary and lower secondary grades are located.

The questionnaire had the following format: every student (or teacher) was asked to fill out every contact they meet at a particular place (e.g. classroom or lunch, cf. Table A4). They also had to indicate how often they usually meet the person. For students, contacts with students from the same class and from a different class occurred at different layers. Every student or teacher was also asked which hobby activities they attend or teach, respectively. The frequency of contacts was identified by different options in the questionnaire: several times a day, once per day, every day, sometimes, rarely. Answers differed between types of contact (e.g. every day for lunch, but once per day for break). Refer to sections A4 and A5 for complete transcripts of electronic questionnaires.

We have also used a summarized timetable that contains data of how many hours per week a teacher has in each class. Using this information, we generated edges between a teacher and all students in a class where they teach, proportionally weighted by the number of hours per week.

The contact networks were constructed from the questionnaire data as follows. The reported contacts are directed (a respondent report whom they are in contact with), but the underlying transmission network is undirected, we symmetrized the responses by the maximum value in each dyad. This also helps with imputing missing responses, as the response rate among pupils was 63% and among teachers 23%, and thus some sort of imputation of missing contacts was necessary. The social networks of pupils and teachers are presented in Table A1, two nodes in the graph are connected whenever the contact between them exists at least in one of the layers.

**Figure A1:**
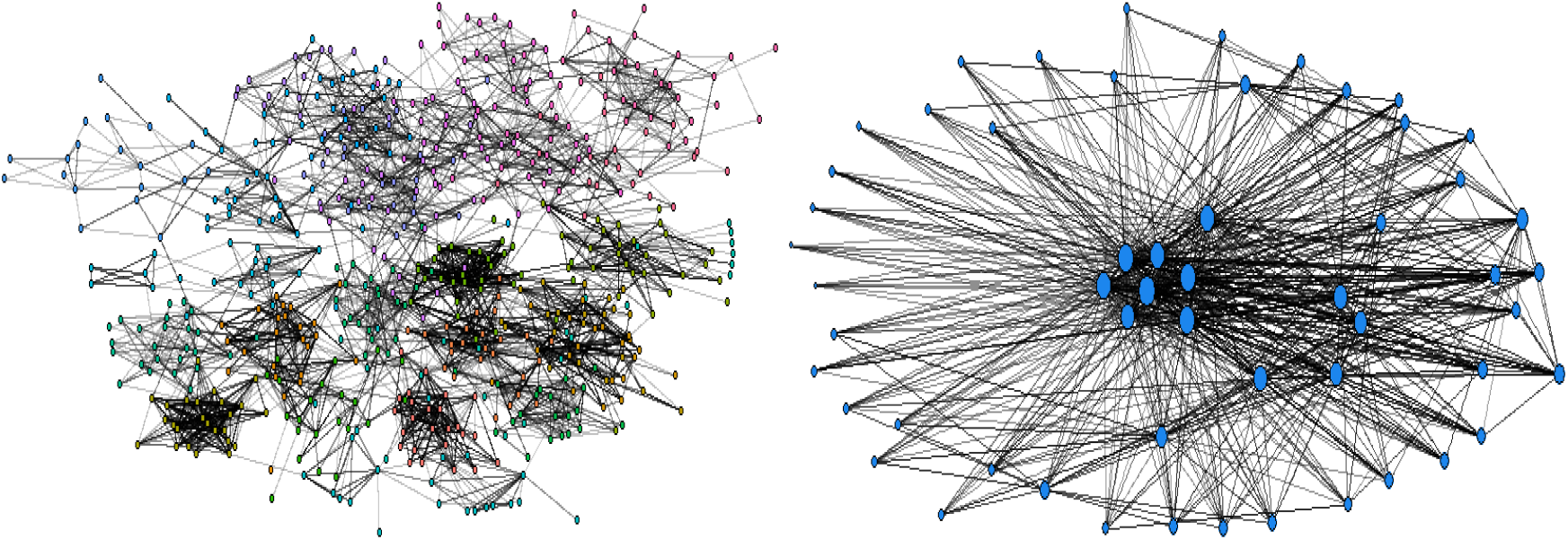
The social network of pupils (left) and teachers (right).

Table A3 presents comparison of graph densities for contacts among three main subsets of our graph — teachers, primary and secondary pupils. It can be seen that teachers contact network is very dense in comparison to the rest of contacts. Secondary pupils have about 1.5 times more contacts than primary pupils, while the contacts between younger and older pupils are relatively sparse.

**Table A1:** Pupil-to-pupil contact networks – descriptive measures

|  | after-school care | desk | outside | lunch | break |
| --- | --- | --- | --- | --- | --- |
| Nodes | 624 | 624 | 624 | 624 | 624 |
| Edges | 1558 | 1018 | 2356 | 3096 | 9364 |
| Avg Degree | 2.497 | 1.631 | 3.776 | 4.962 | 15.006 |
| Centralization | 0.038 | 0.009 | 0.034 | 0.031 | 0.035 |
| Density | 0.008 | 0.005 | 0.012 | 0.016 | 0.048 |
| Connectedness | 0.012 | 0.015 | 0.746 | 0.028 | 1 |
| Closure | 0.553 | 0.235 | 0.225 | 0.490 | 0.663 |
| Avg Distance | 1.841 | 3.450 | 6.017 | 1.990 | 4.790 |

#### Building a contact network

After the data preprocessing, we have obtained a network with 624 student nodes, 55 teacher nodes, and 27,677 edges. The edges are divided into 98 layers corresponding to four types of contacts: students in class, teacher in class, after-school clubs, and others (friendship, more contacts). The overview of layers is presented in Table A4.

**Table A2:** Teacher-to-teacher contact networks – descriptive measures

|  | office | school | outside |
| --- | --- | --- | --- |
| Nodes | 55 | 55 | 55 |
| Edges | 1122 | 50 | 458 |
| Avg Degree | 20.400 | 0.909 | 8.327 |
| Centralization | 0.646 | 0.136 | 0.878 |
| Density | 0.378 | 0.017 | 0.154 |
| Connectedness | 1 | 0.145 | 1 |
| Closure | 0.505 | 0 | 0.251 |
| Avg Distance | 1.622 | 3.740 | 1.846 |

**Table A3:**
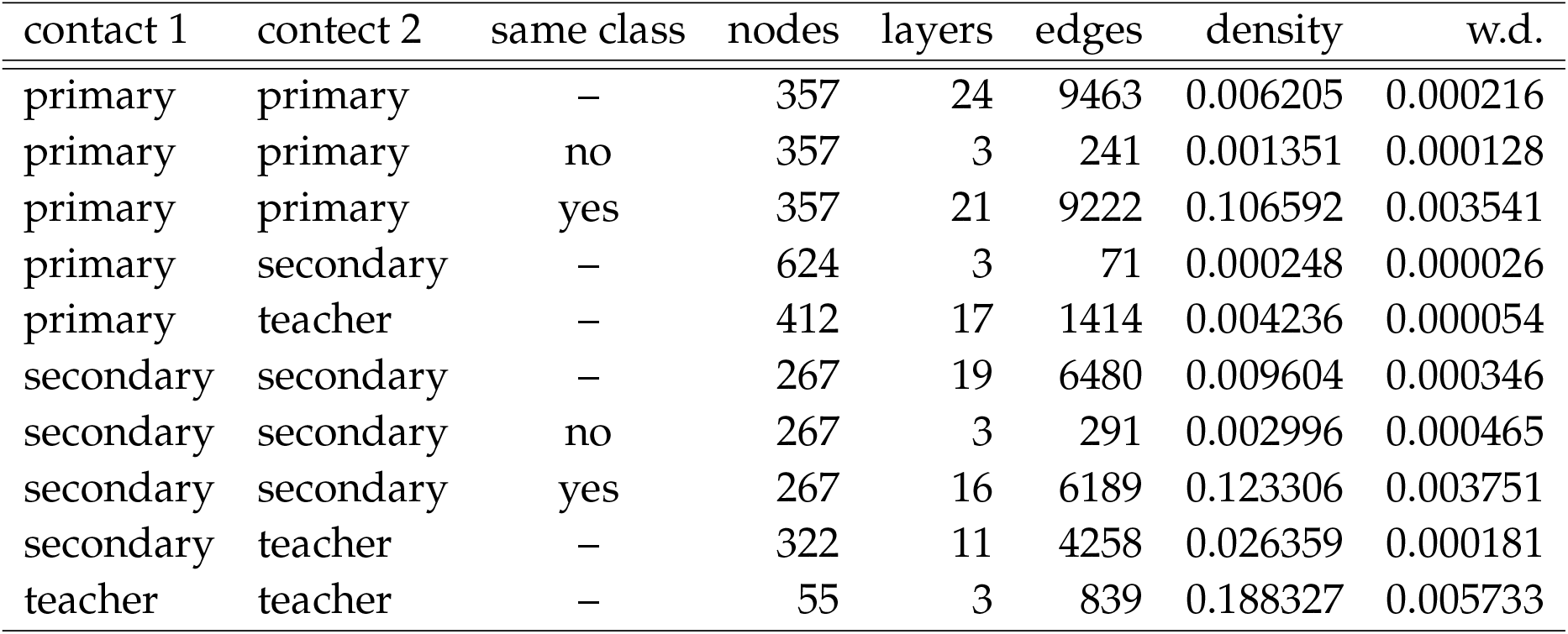
Subgraph densities for interactions among three categories of nodes – teachers, primary and secondary pupils. For pupils we further differentiate contacts inside and outside of classes. Weighted density (w.d.) represents a value where contacts are weighted by their intensity and weight of the layer.

The frequencies of contacts and their types are then used to assess contact intensities. The intensities of contact types are taken from our epidemic model of the city, and they were obtained by survey of experts processed by the Saaty method. The contact intensity values used in our school model are presented in Table A5. For details, cf. Berec et al. (2021). The assignment of intensities and their combinations to layer types is presented in column two of Table A4.

Figure A2 and Table A6 show the results of a simulation of infection spread in a fully open school (no infection control measures) identifying the sources of infections. The majority of infections (43%) originate in class and during lunch (19%). Teachers are subject of two types of interactions (teacher-teacher and teacher-student) which together result in 17% of infections.

**Figure A2:**
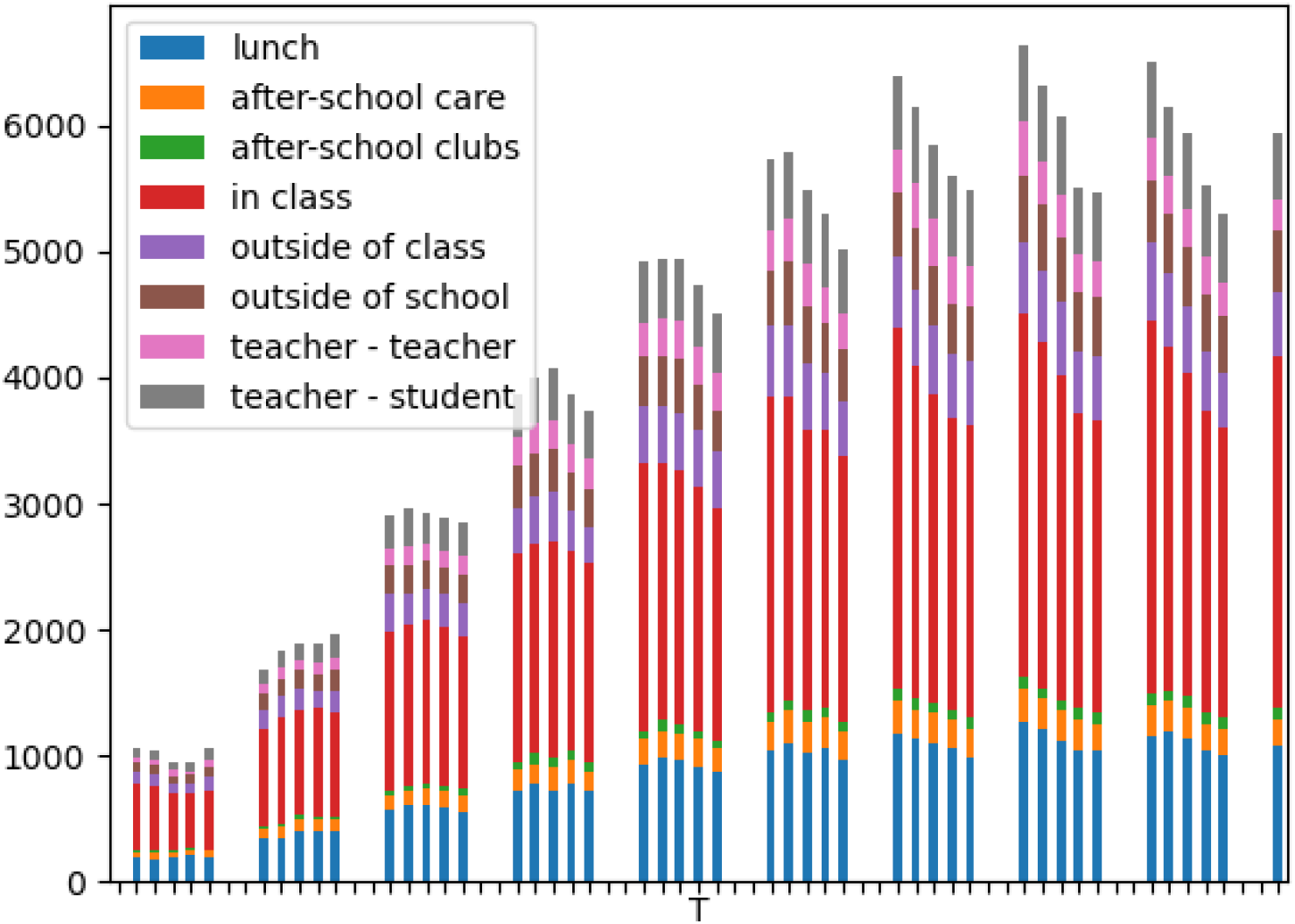
Sources of infections in open school baseline scenario. The graph represents daily values of infected individuals categorized by the contact type. Numbers are sums over 1000 simulation runs. Average numbers in percent are presented in Table A6.

**Table A4:** Overview of the types of social contact layers. The contact intensity column refers to basic contact intensities from Table A5.

| layer | contact intensity | note |
| --- | --- | --- |
| break | closerandomcontact |  |
| lunch | dininingcontact |  |
| after_school_care | closerandomcontact | Only some students go there |
| desk-mate_main | closelongtermcontact | Respondent sits at the same desk with this person the most frequently (in Czechia, students usually sit in pairs) |
| outside_of_school | $(\text{closelongtermcontact} + \text{closeopenair} + \text{closerandomcontact})/3$ | Any contact when not at school |
| outside_of_school_other | $(\text{closelongtermcontact} + \text{closeopenair} + \text{closerandomcontact})/4$ | |
| desk-mate_sometimes | closelongtermcontact | Respondent sometimes sits at the same desk with this person |
| break_other | closelongtermcontact |  |
| desk-mate_sometimes_other | $(\text{closelongtermcontact} + \text{closerandomcontact})/2$ | |
| office | $(\text{closerandomcontact} * 4 + \text{closelongtermcontact})/5$ | These teachers share an office |
| at_job | closerandomcontact |  |
| outside_job | $(\text{closelongtermcontact} + \text{closeopenair} + \text{closerandomcontact})/3$ | Any contact when not at school |
| teaches_class_XY (27 classes) | distantlongtermcontact | Teacher x student contact |
| Assistent_class_XY (2 classes) | closelongtermcontact | Teacher x student contact |
| hobby (26) | closerandomcontact | Student x student contact |
| active_hobby (5) | physicalshortterm | Student x student contact |
| class_contact (27) | closerandomcontact | Student x student contact |

**Table A5:** Contact categories and their intensities.

| contact name | intensity |
| --- | --- |
| closelongtermcontact | 0.255905135951299 |
| physicallongterm | 0.209577963668343 |
| physicalshortterm | 0.209577963668343 |
| dininingcontact | 0.156907515722802 |
| distantlongtermcontact | 0.108580705683382 |
| closerandomcontact | 0.031870115278543 |
| serviceclientcontact | 0.017146327621734 |
| closeopenair | 0.010434272405554 |

**Table A6:** Percentage of sources of infections over layers in open school baseline scenario.

|  |  |  |  |
| --- | --- | --- | --- |
| lunch | 19.01 % | outside of class | 9.11 % |
| after-school care | 4.19 % | outside of school | 7.99 % |
| after-school clubs | 1.53 % | teacher - teacher | 5.56 % |
| in class | 42.96 % | teacher - student | 9.64 % |

### A3 Results

In the following we present complete results of our three experiments that compare relative efficiency of the following measures: weekly rotations, testing, and their combination. Furthermore, we present an experiment to estimate the importance of teachers in the epidemics spread within school. For each experiment we present the results in four epidemics levels represented by average import of infected individuals. Table A7 contains numbers of individuals relative to 100 thousand inhabitants. The levels were selected to represent severe epidemics and its ratios of 50%, 25% and 10%.

**Table A7:** Levels of epidemics used in our simulations. Numbers of infected calculated per 100 thousand inhabitants daily and weekly.

| Epidemics | daily import per 100k | weekly import per 100k |
| --- | --- | --- |
| 100% – severe | 146.38 | 1024.69 |
| 50% – medium | 73.66 | 515.61 |
| 25% – quarter | 36.36 | 254.54 |
| 10% – weak | 14.64 | 102.47 |

#### Rotations

**Table A8:**
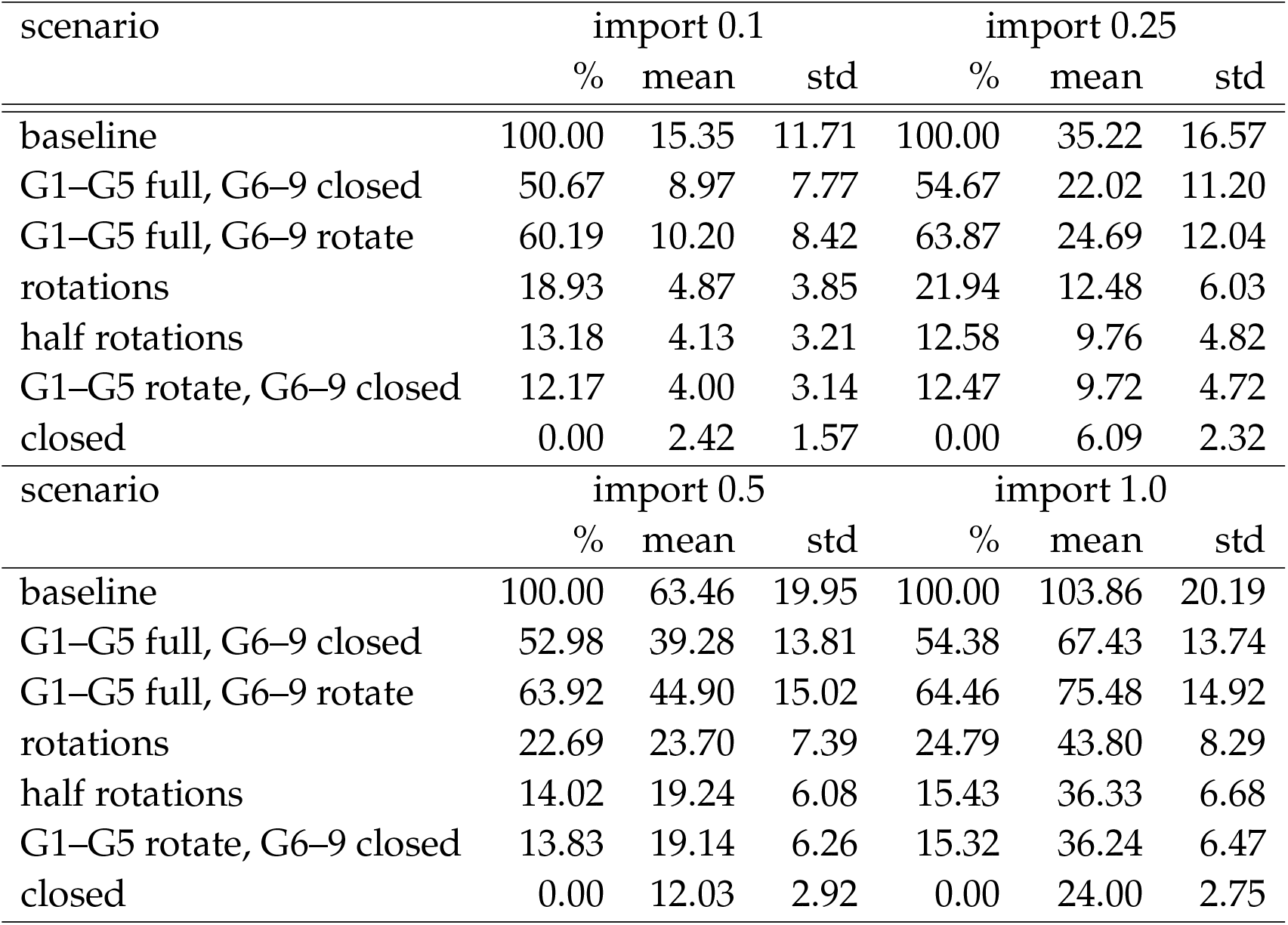
Rotation scenarios. Percentage of numbers of infected six weeks after the start of the policy.

| scenario | import 0.1 |  |  | import 0.25 |  |  |
| --- | --- | --- | --- | --- | --- | --- |
|  | % | mean | std | % | mean | std |
| baseline | 100.00 | 15.35 | 11.71 | 100.00 | 35.22 | 16.57 |
| G1–G5 full, G6–9 closed | 50.67 | 8.97 | 7.77 | 54.67 | 22.02 | 11.20 |
| G1–G5 full, G6–9 rotate | 60.19 | 10.20 | 8.42 | 63.87 | 24.69 | 12.04 |
| rotations | 18.93 | 4.87 | 3.85 | 21.94 | 12.48 | 6.03 |
| half rotations | 13.18 | 4.13 | 3.21 | 12.58 | 9.76 | 4.82 |
| G1–G5 rotate, G6–9 closed | 12.17 | 4.00 | 3.14 | 12.47 | 9.72 | 4.72 |
| closed | 0.00 | 2.42 | 1.57 | 0.00 | 6.09 | 2.32 |
| scenario | import 0.5 |  |  | import 1.0 |  |  |
|  | % | mean | std | % | mean | std |
| baseline | 100.00 | 63.46 | 19.95 | 100.00 | 103.86 | 20.19 |
| G1–G5 full, G6–9 closed | 52.98 | 39.28 | 13.81 | 54.38 | 67.43 | 13.74 |
| G1–G5 full, G6–9 rotate | 63.92 | 44.90 | 15.02 | 64.46 | 75.48 | 14.92 |
| rotations | 22.69 | 23.70 | 7.39 | 24.79 | 43.80 | 8.29 |
| half rotations | 14.02 | 19.24 | 6.08 | 15.43 | 36.33 | 6.68 |
| G1–G5 rotate, G6–9 closed | 13.83 | 19.14 | 6.26 | 15.32 | 36.24 | 6.47 |
| closed | 0.00 | 12.03 | 2.92 | 0.00 | 24.00 | 2.75 |

**Figure A3:**
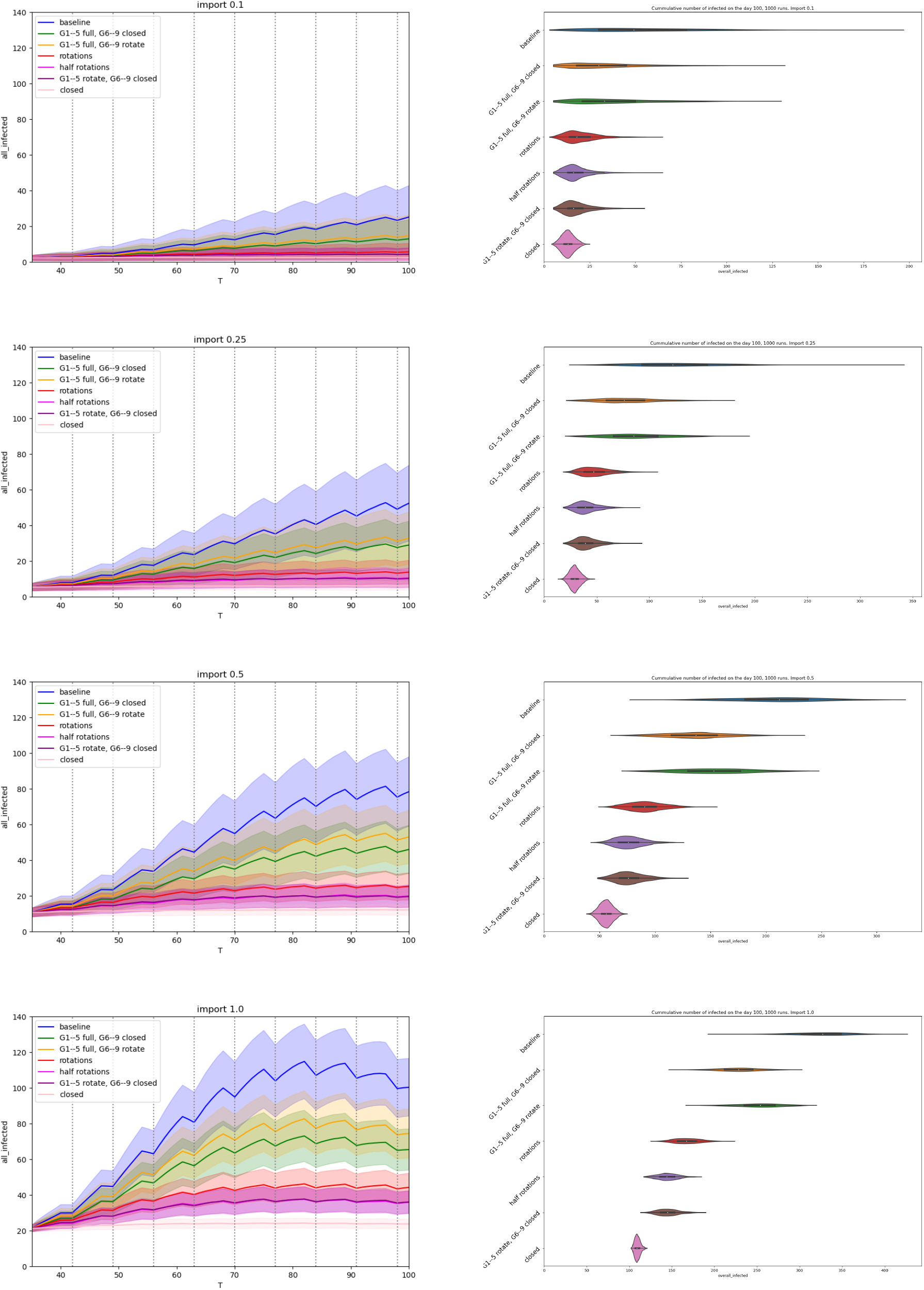
Rotation scenarios. (left) Comparison of number of infected active cases during the run. The x-axis represents days of simulation, the y-axis represents number of infections at school, mean values with standard deviation are plotted for each simulation. (right) Violin plots: all infected (cumulative over the whole run).

#### Testing

**Table A9:** Testing scenarios. Percentage of numbers of infected six weeks after the start of the policy. The numbers in brackets denote the sensitivity of the test.

| scenario | import 0.1 |  |  | import 0.25 |  |  |
| --- | --- | --- | --- | --- | --- | --- |
|  | % | mean | std | % | mean | std |
| baseline | 100.00 | 15.35 | 11.71 | 100.00 | 35.22 | 16.57 |
| PCR test once | 38.34 | 7.38 | 6.59 | 42.26 | 18.40 | 10.34 |
| antigen test twice (0.1) | 83.43 | 13.21 | 11.05 | 83.28 | 30.35 | 14.33 |
| antigen test twice (0.2) | 64.02 | 10.70 | 8.96 | 67.91 | 25.87 | 13.24 |
| antigen test twice (0.4) | 43.48 | 8.04 | 6.82 | 44.42 | 19.03 | 10.31 |
| antigen test once (0.1) | 90.87 | 14.17 | 11.29 | 93.36 | 33.28 | 16.12 |
| antigen test once (0.2) | 79.34 | 12.68 | 10.36 | 82.02 | 29.98 | 14.89 |
| antigen test once (0.4) | 61.27 | 10.34 | 8.61 | 64.29 | 24.82 | 12.65 |
| closed | 0.00 | 2.42 | 1.57 | 0.00 | 6.09 | 2.32 |
|  | import 0.5 |  |  | import 1.0 |  |  |
|  | % | mean | std | % | mean | std |
| baseline | 100.00 | 63.46 | 19.95 | 100.00 | 103.86 | 20.19 |
| PCR test once | 42.26 | 33.77 | 12.18 | 44.59 | 59.62 | 13.44 |
| antigen test twice (0.1) | 83.60 | 55.03 | 17.73 | 85.27 | 92.10 | 18.02 |
| antigen test twice (0.2) | 67.55 | 46.77 | 15.67 | 71.33 | 80.97 | 16.59 |
| antigen test twice (0.4) | 45.98 | 35.68 | 12.25 | 48.43 | 62.68 | 13.34 |
| antigen test once (0.1) | 91.26 | 58.97 | 19.18 | 92.43 | 97.82 | 18.81 |
| antigen test once (0.2) | 82.13 | 54.27 | 18.06 | 83.74 | 90.88 | 18.58 |
| antigen test once (0.4) | 65.82 | 45.88 | 15.57 | 68.77 | 78.92 | 16.99 |
| closed | 0.00 | 12.03 | 2.92 | 0.00 | 24.00 | 2.75 |

**Figure A4:**
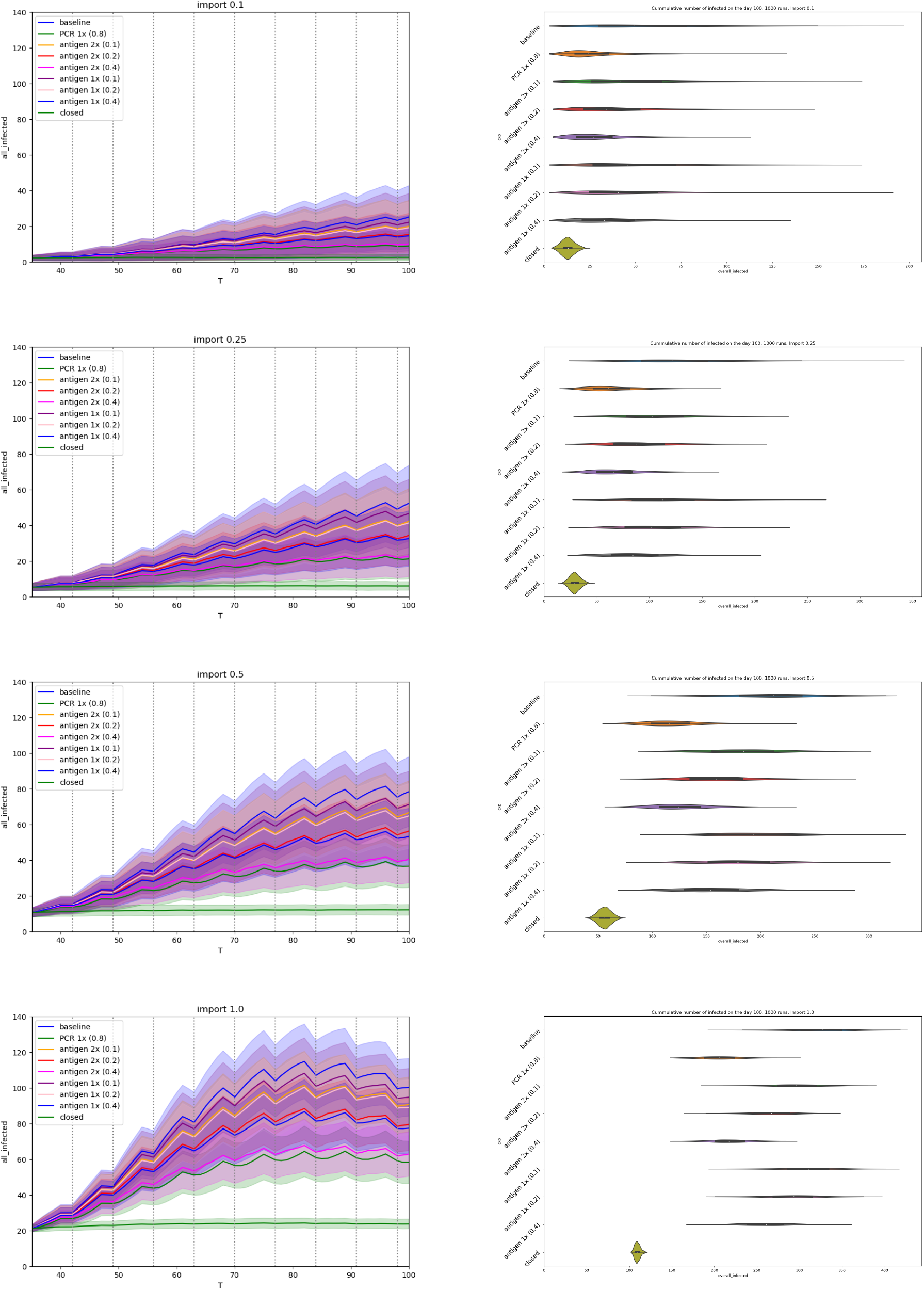
Testing scenarios. (left) Comparison of number of infected active cases during the run. The x-axis represents days of simulation, the y-axis represents number of infections at school, mean values with standard deviation are plotted for each simulation. (right) Violin plots: all infected (cumulative over the whole run).

#### Rotations and testing

**Table A10:**
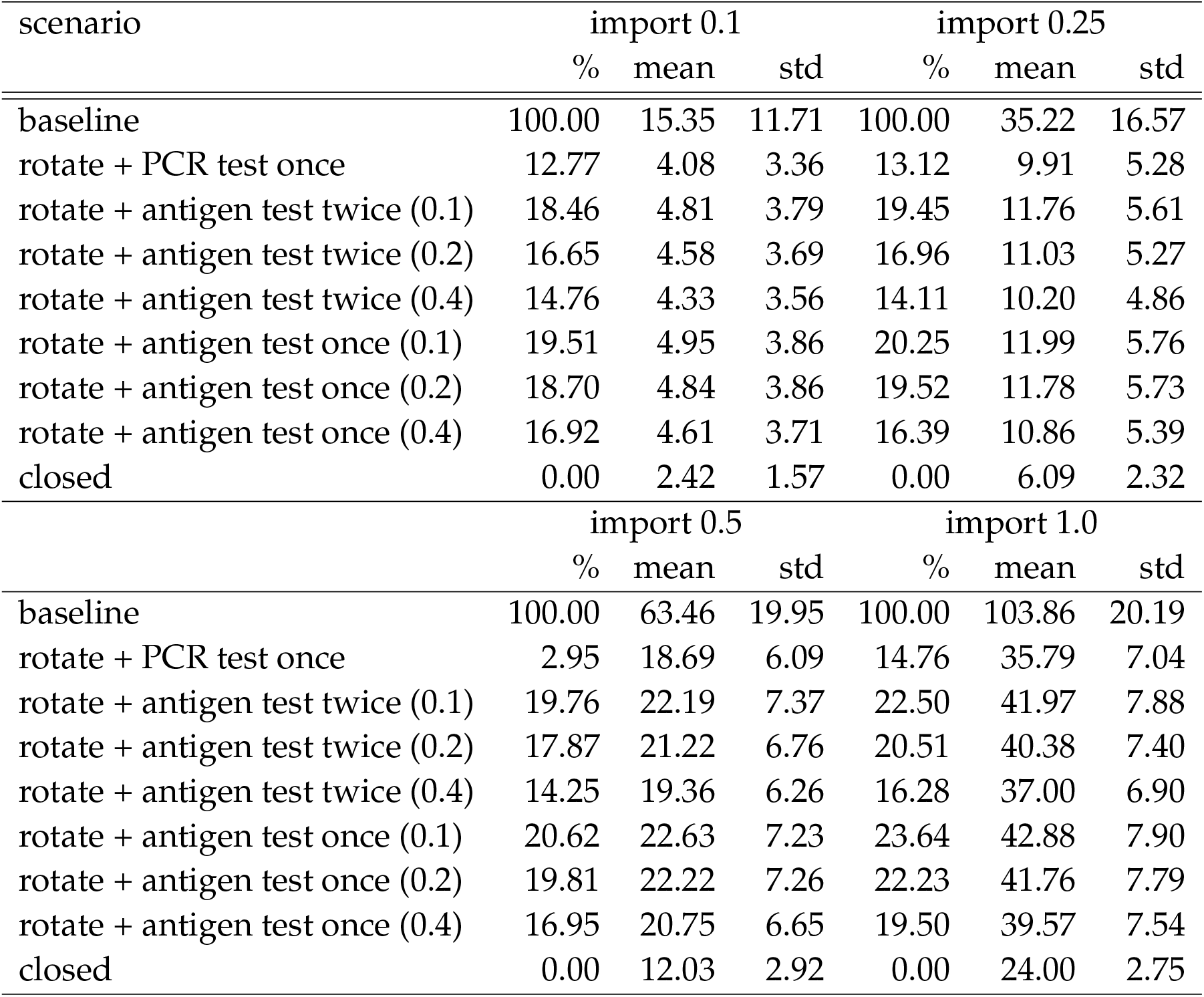
Rotation and testing scenarios. Percentage of numbers of infected six weeks after the start of the policy.

**Figure A5:**
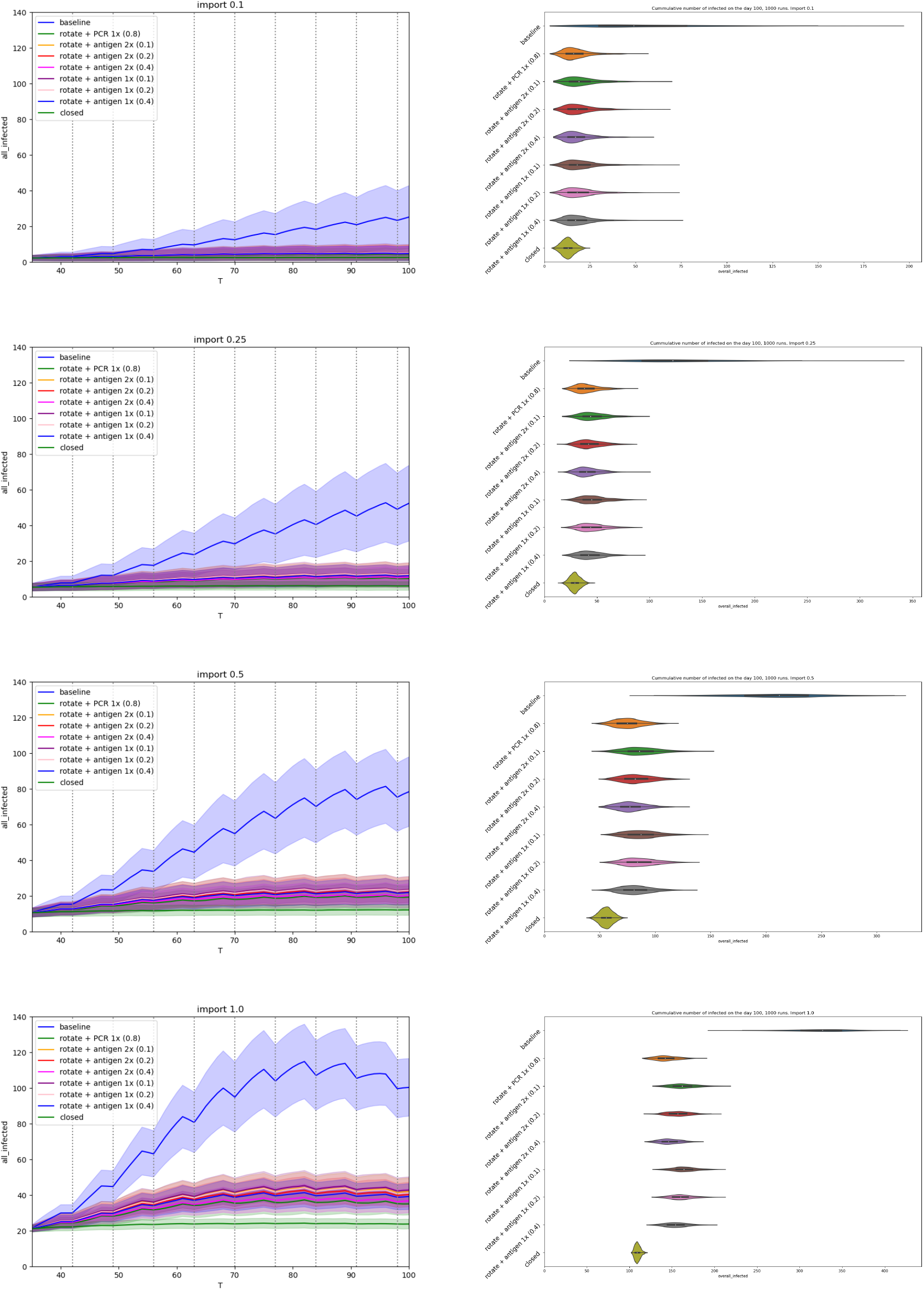
Rotating + testing scenarios. (left) Comparison of number of infected active cases during the run. The x-axis represents days of simulation, the y-axis represents number of infections at school, mean values with standard deviation are plotted for each simulation. (right) Violin plots: all infected (cumulative over the whole run).

#### The role of teachers

**Table A11:**
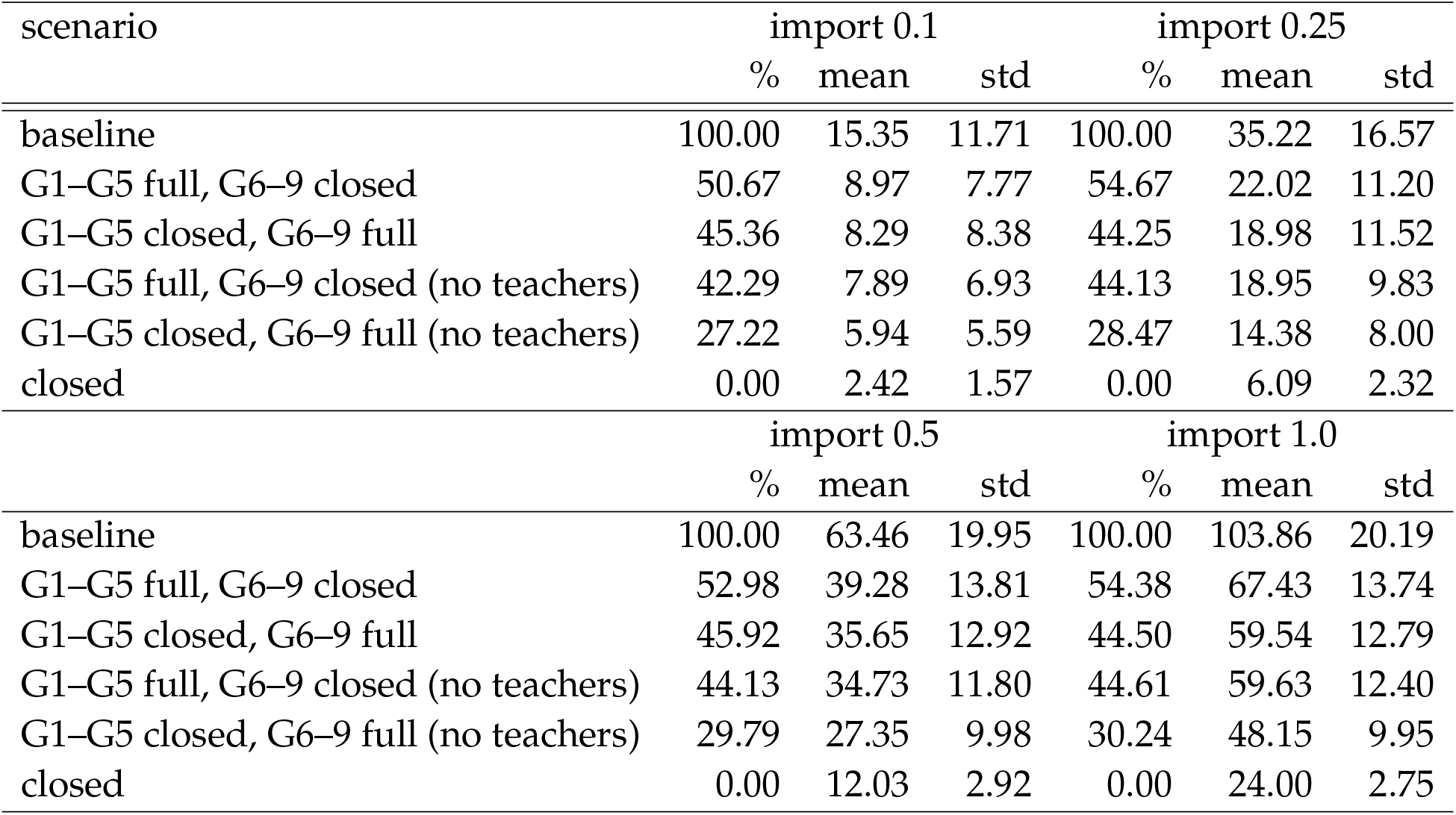
The role of teachers. Percentage of numbers of infected six weeks after the start of the policy.

**Figure A6:**
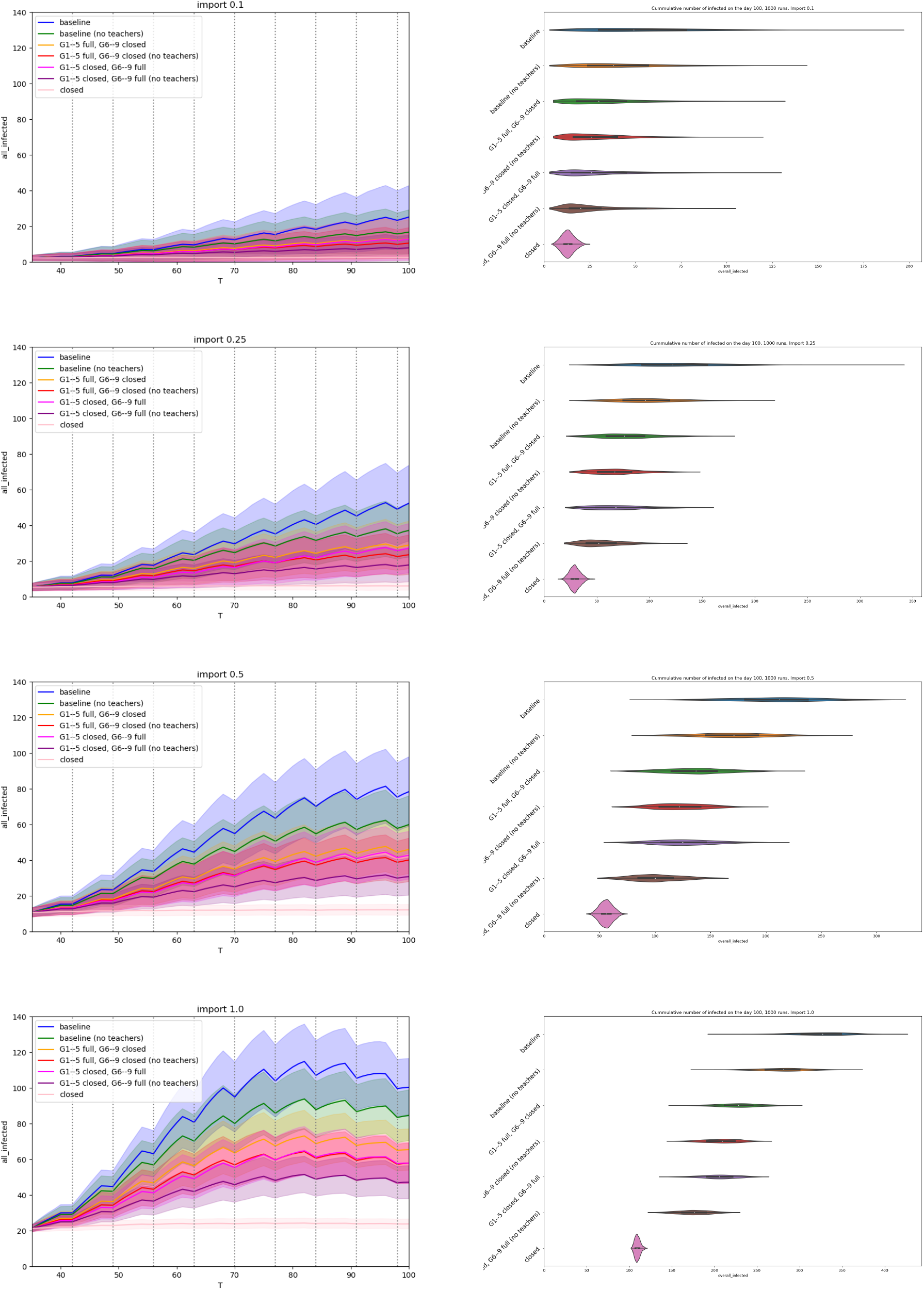
The role of teachers. (left) Comparison of number of infected active cases during the run. The x-axis represents days of simulation, the y-axis represents number of infections at school, mean values with standard deviation are plotted for each simulation. (right) Violin plots: all infected (cumulative over the whole run).

### A4 Questionnaire students

Students filled in the questionnaires electronically. Parents of children in Grades 1-3 were instructed to help children filling in it. A transcription of the online version of the questionnaire follows.

#### Introduction

Dear students,

Completing this questionnaire will help to find out which measures in schools are the most effective against the spread of coronavirus. Therefore, we ask you to think about the questions and fill them in truthfully. Every answer matters. Careful completion of the questionnaire should not take more than 10 minutes.

You cannot return to the questionnaire, so please fill in the questionnaire at once. In the questionnaire, you will fill in who you meet at school and out of school. You will fill in personal information such as name, but it will remain safe. Only one designated school staff member will work with the personal data, converting all the names to anonymous codes and then deleting the names. No one from the research team, no other school staff, or your classmates will know how you answered this questionnaire.

The questions refer to the standard times, i.e. without school closure or restrictions of other activities. The goal is to find out who you normally talk to and who you meet.

Research team

Ing. René Levinský, Ph.D.

Center for modeling biological and social processes

#### Personal information

What is your name?

What grade and class do you attend?

#### Participation in school activities

Do you regularly attend after school club? YES NO

Do you regularly have lunch in the school cafeteria? YES NO

#### School breaks

*The names of students in particular classes was filled in by a designated school employee and was not accessible by researchers*.

With whom <u>from your class</u> do you usually spend time <u>during breaks</u>?

*Your name is also in the list, there please fill in “Several times a day”*.

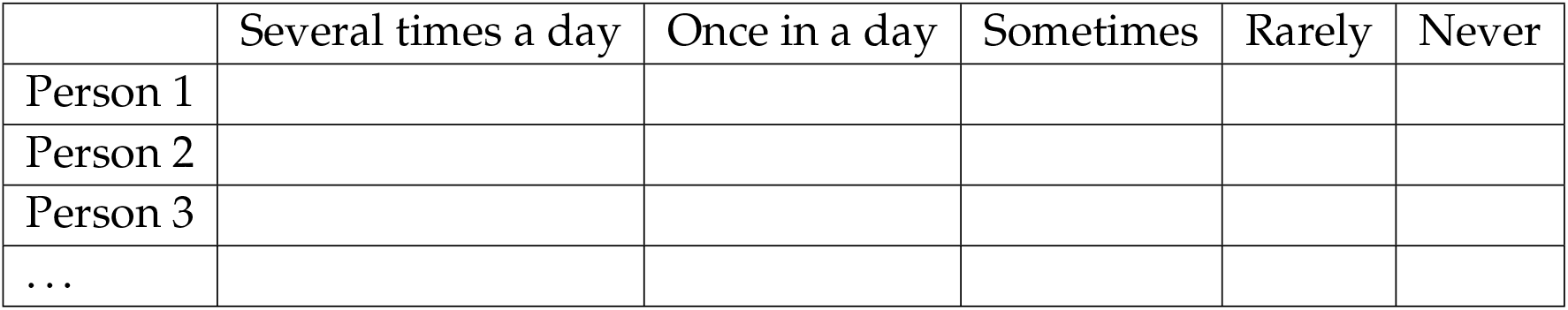

With whom outside of your class do you usually spend time <u>during breaks</u>?

*Please fill in their names, grade and choose frequency how often do you meet with them. The maximum number of contacts is 10*.

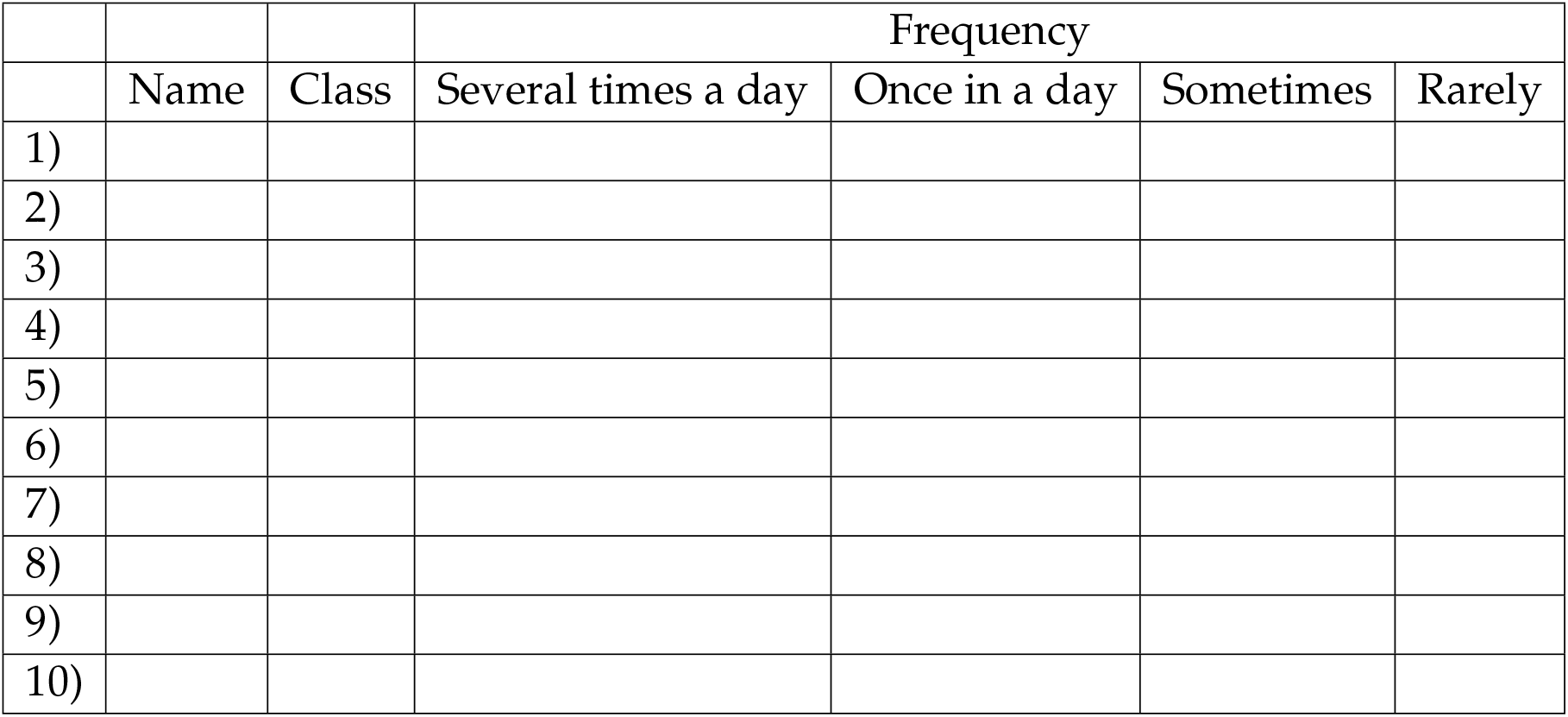

#### Outside of school

With whom <u>from your class</u> do you usually spend time outside of school (e.g. on the way to school, during your free time, in sports activities)?

*First check people, you spend time with outside of school and then fill in frequency. Your name is also on the list, there do not check the box*.

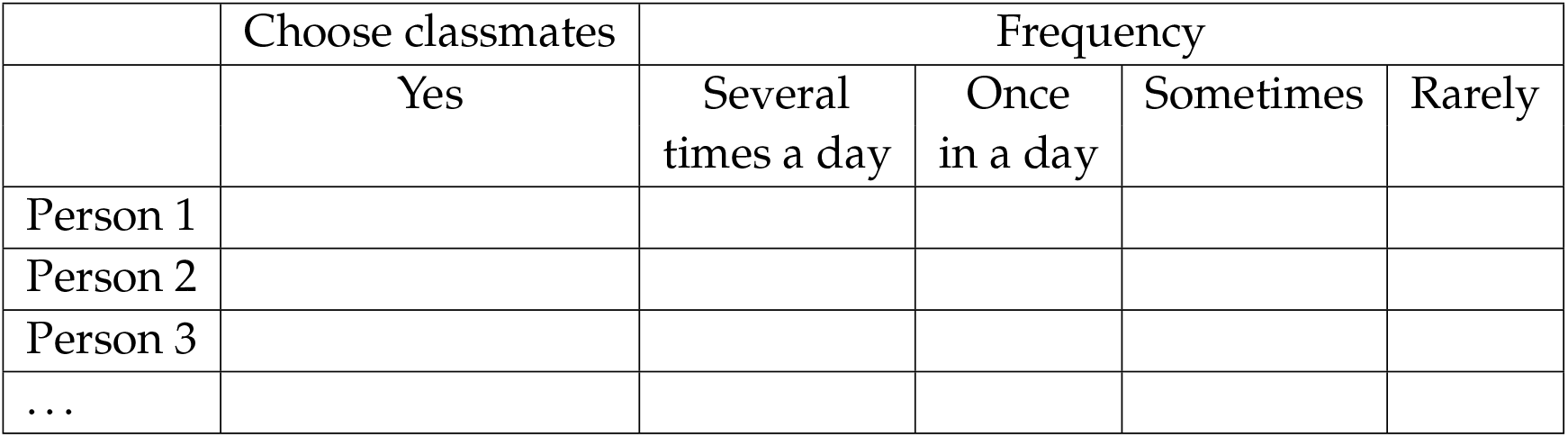

With whom outside of your class do you usually spend time outside of school (e.g. on the way to school, during your free time, in sports activities)?

*Please fill in their names, grade and choose frequency how often do you meet with them. The maximum number of contacts is 10*.

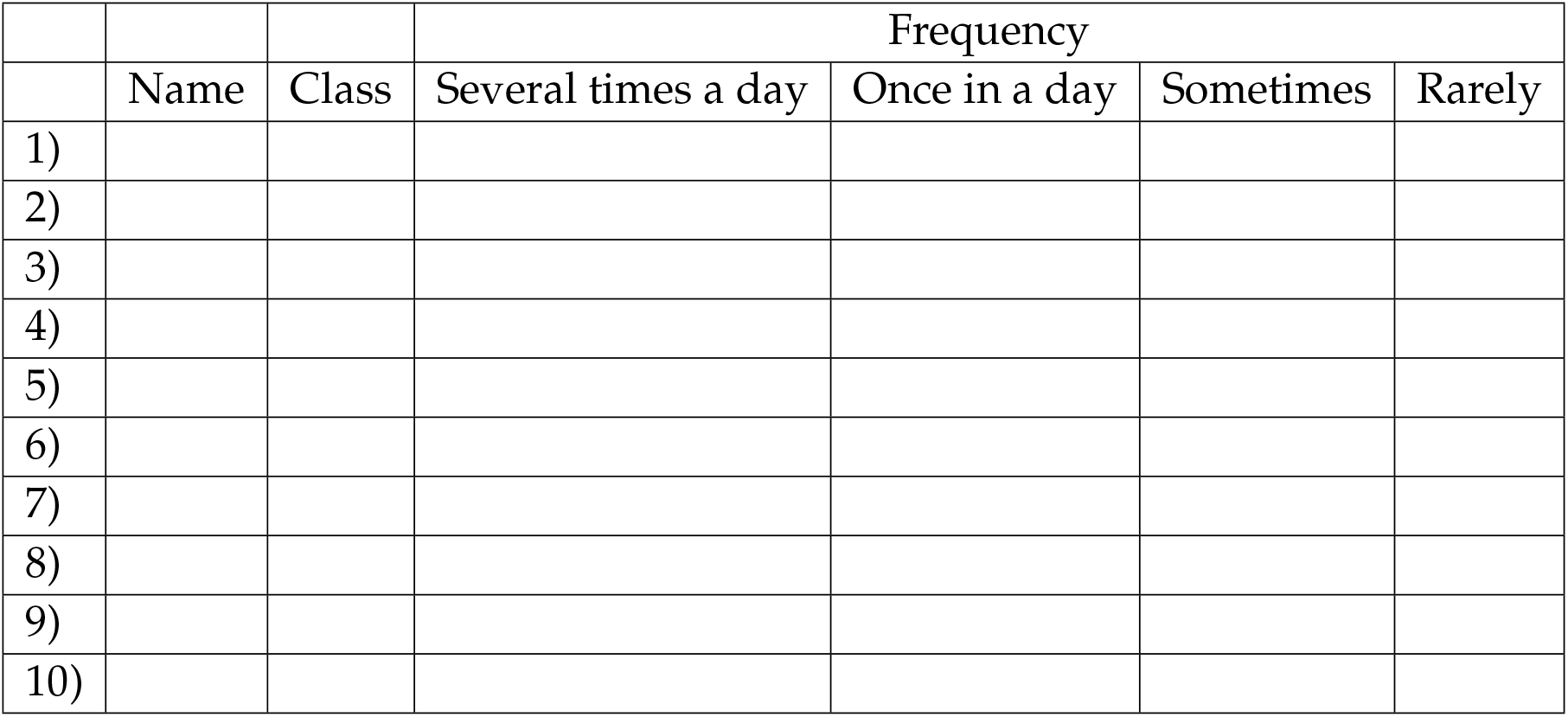

#### Lunch

*Filter for students who regularly have lunch in the school cafeteria*

With whom <u>from your class</u> do you usually spend time during lunch?

*First check people, you spend time with during lunch and then fill in frequency. Your name is also on the list, there do not check the box*.

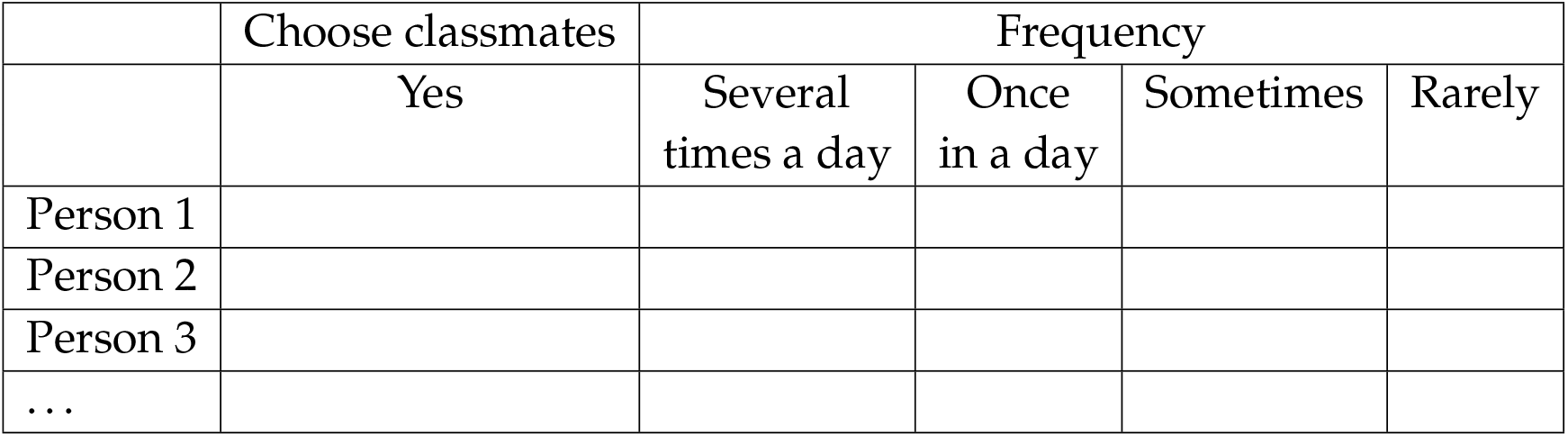

With whom <u>outside of your class</u> do you usually spend time <u>during lunch</u>?

*Please fill in their names, grade and choose frequency how often do you meet with them. The maximum number of contacts is 10*.

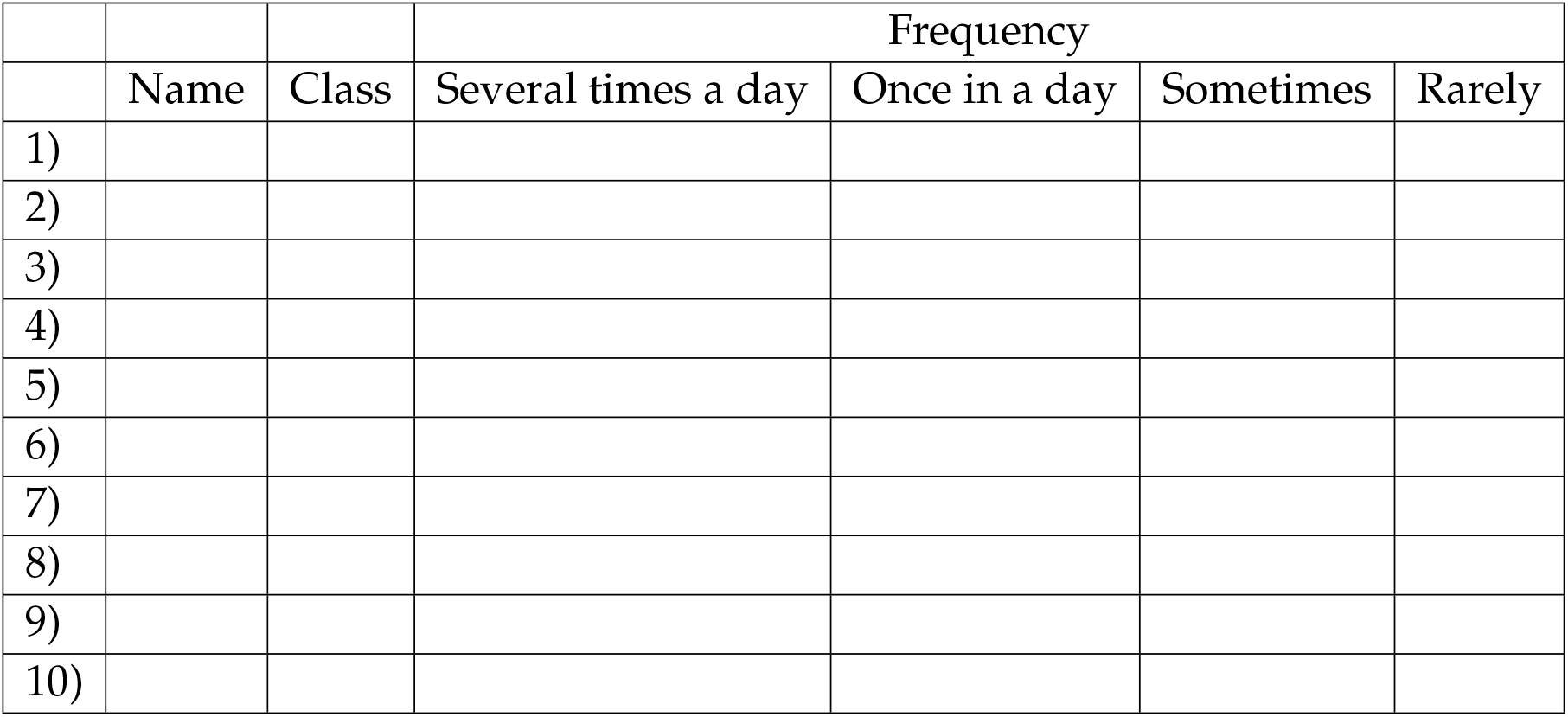

#### After school club

*Filter for students who regularly attend afterschool club*

With whom <u>from your class</u> do you usually spend time in the after school club?

*First check people, you spend time in the after school club and then fill in frequency. Your name is also on the list, there do not check the box*.

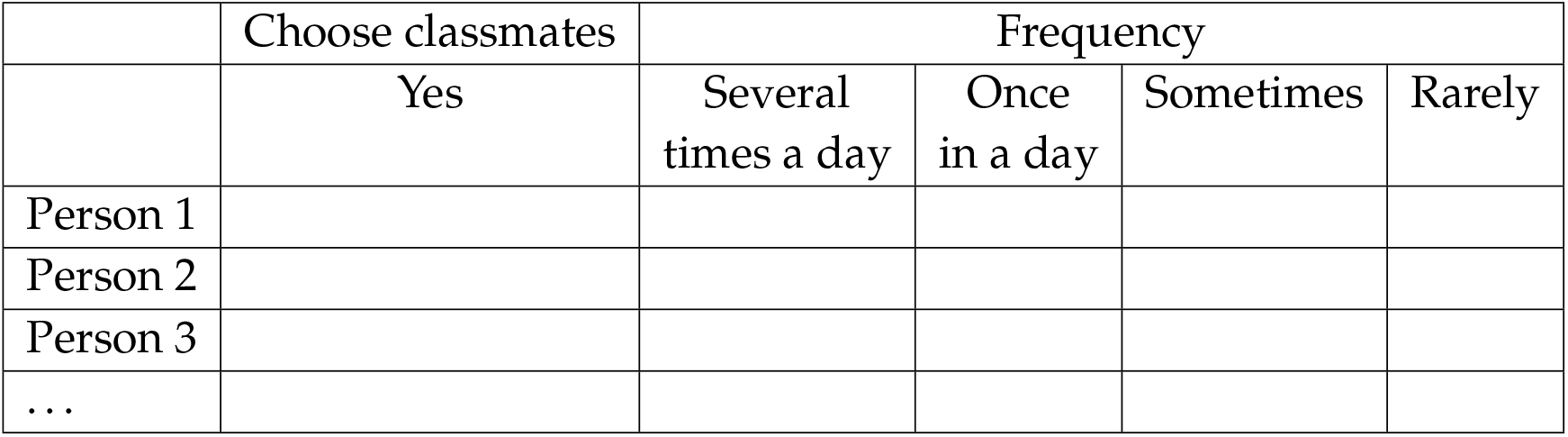

With whom outside of your class do you usually spend time in the after school club?

*Please fill in their names, grade and choose frequency how often do you meet with them. The maximum number of contacts is 10*.

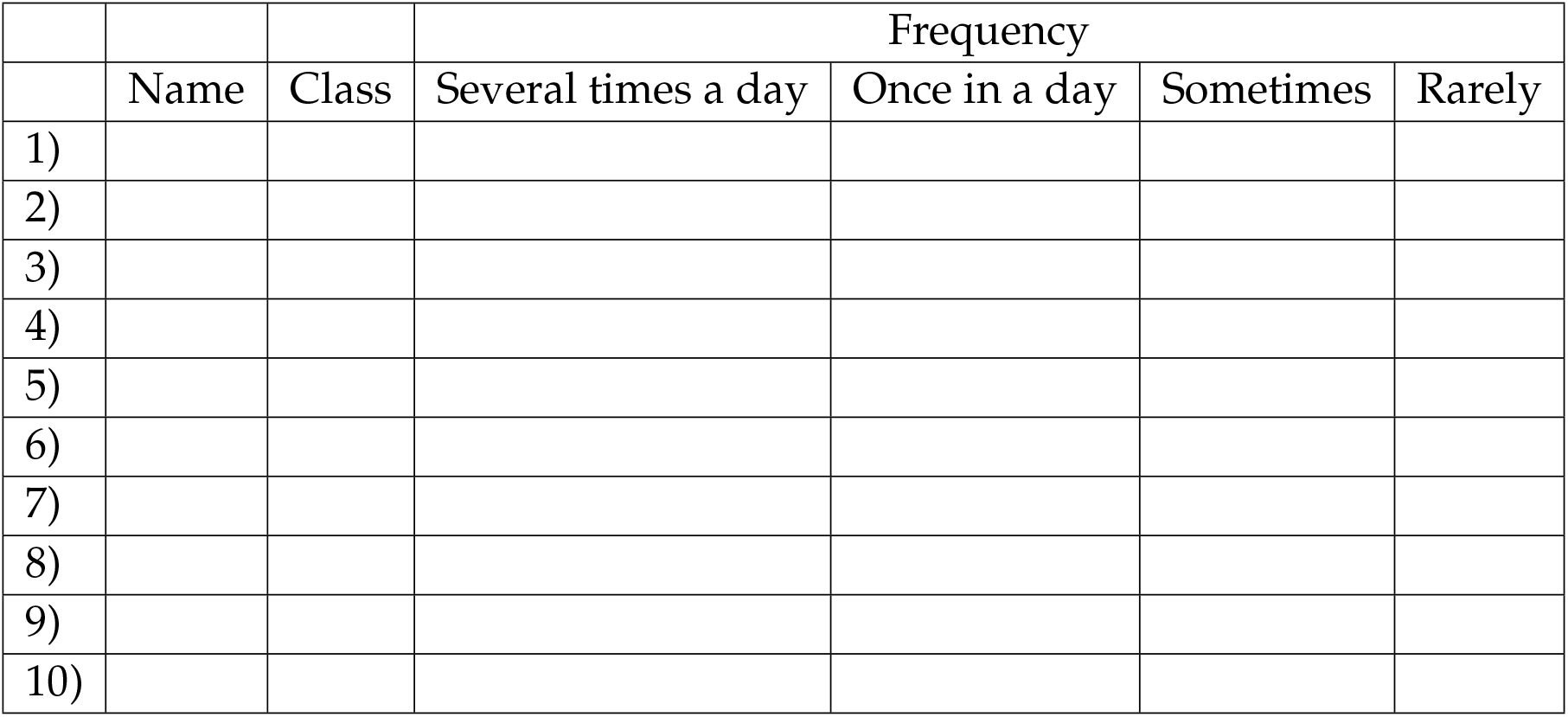

With whom do you regularly sit next at your school desk?

*Fill in the name. In case you sit alone, fill in “alone”*.

Do you sit next to someone else <u>from your class</u> during some lessons?

*Fill in name of a maximum of five classmates*.

1)

2)

3)

4)

5)

Do you sit next to someone else from another class during some lessons?

*Fill in name of a maximum of three classmates*.

1)

2)

3)

Do you attend any of the school free time activities?

*Check all that apply*.

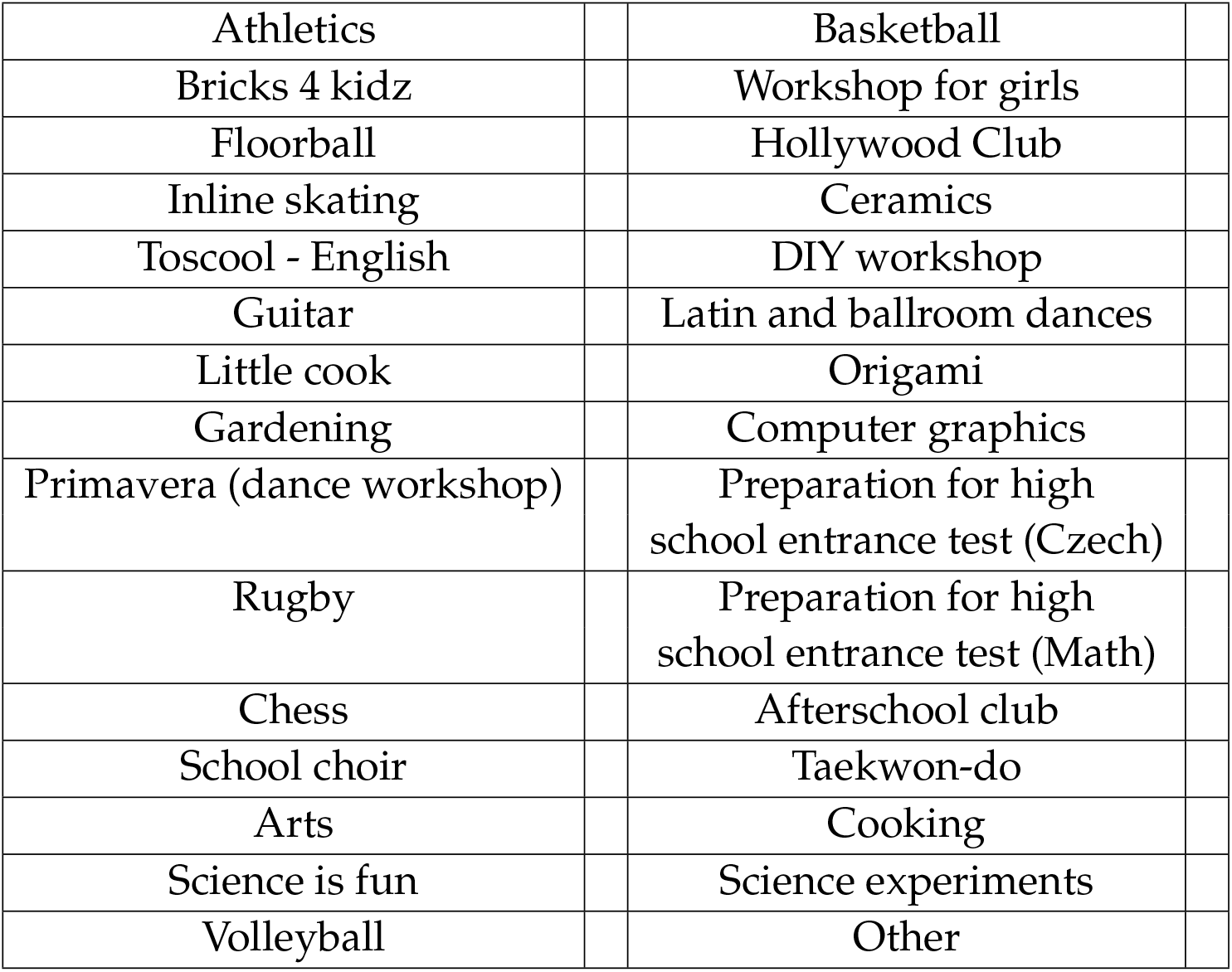

|  |  |  |
| --- | --- | --- |
| Athletics |  | Basketball |
| Bricks 4 kidz |  | Workshop for girls |
| Floorball |  | Hollywood Club |
| Inline skating |  | Ceramics |
| Toscool - English |  | DIY workshop |
| Guitar |  | Latin and ballroom dances |
| Little cook |  | Origami |
| Gardening |  | Computer graphics |
| Primavera (dance workshop) |  | Preparation for high school entrance test (Czech) |
| Rugby |  | Preparation for high school entrance test (Math) |
| Chess |  | Afterschool club |
| School choir |  | Taekwon-do |
| Arts |  | Cooking |
| Science is fun |  | Science experiments |
| Volleyball |  | Other |

### A5 Questionnaire teachers

Teachers filled in the questionnaires electronically. This is a transcription of the online version.

#### Introduction

Dear school employees,

The second wave of the coronavirus pandemic led to temporary school closures. The Ministry of Education, in cooperation with the academic community, is now looking for ways to open schools safely as soon as possible. By completing this questionnaire, you will provide data to help model the effects of various measures on the spread of coronavirus in schools (e.g. grade rotation scheme, afterschool club and school cafeteria closure, or class division and rotation scheme). It should take no more than 15 minutes to complete the questionnaire. You cannot return to the questionnaire, so please fill in the questionnaire at once.

You will fill in personal data in the questionnaire, but it will remain safe. Only one member of the research team will work with the data with personal data, who will convert all personal information into anonymous codes and delete the personal data. Consequently, no one will be able to identify individual answers. The outputs of the project will always be presented only in aggregated form.

The questions refer to the standard times, i.e. without school closure or restrictions of other activities. The aim is to find out the contacts before pandemics.

Research team

Ing. René Levinský, Ph.D.

Center for modeling biological and social processes

I agree with the processing of personal data in this research project.

YES NO

#### Personal and school information

What is your name?

What is your age category?

- 20-29
- 30-39
- 40-49
- 50-59
- 60 and more

What is your position in your school?

- Teacher
- Educational assistant
- After school club assistant

On what school level do you teach?

- Primary (grade 1-5)
- Lower secondary (grade 6-9)
- Both (grade 1-9)

What school subjects do you teach?

*Check all that apply*

|  |  |  |
| --- | --- | --- |
| Active English |  | English |
| Chemistry |  | History |
| French |  | Music |
| Physics |  | ICT |
| Informatics |  | Conversation in English |
| Mathematics |  | German |
| DIY |  | Biology |
| PE |  | Electives |
| Civics |  | Health education |
| Arts |  | Basics of administration |
| Geography |  |  |

Do you teach any of the following after school activities?

*Check all that apply*

|  |  |  |
| --- | --- | --- |
| Athletics |  | Basketball |
| Floorball |  | Origami |
| Gardening |  | Preparation for high school entrance test (Czech) |
| Rugby |  | Preparation for high school entrance test (Math) |
| School choir |  | Cooking |
| Other |  |  |

#### Contacts

*The full list of pedagogues was included in the following questions. We filter the values based on their answers about their position and grade level*

Whom from your colleagues do you meet <u>in your school office</u>?

*In case you do not meet the person in a given row, do not check any value. Your name is also in the list, do not fill any value in that row*.

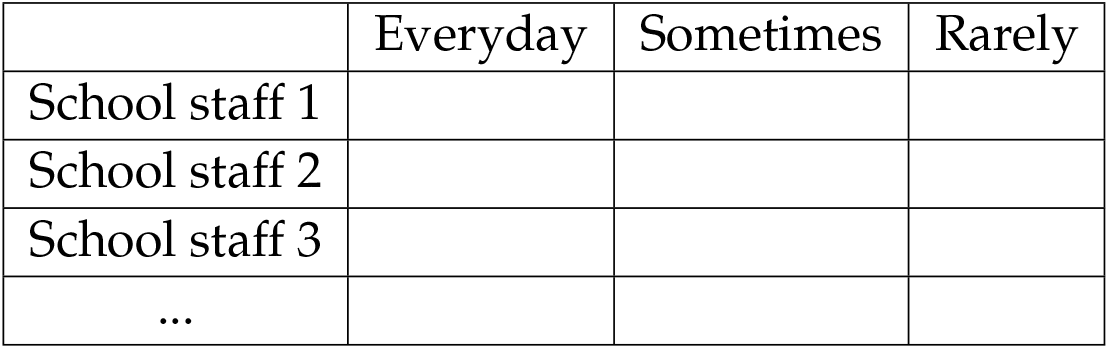

With whom from your colleagues do you spend time in school (e.g. during teaching, breaks, lunch)?

*In case you do not meet the person in a given row, do not check any value. Your name is also in the list, do not fill any value in that row*.

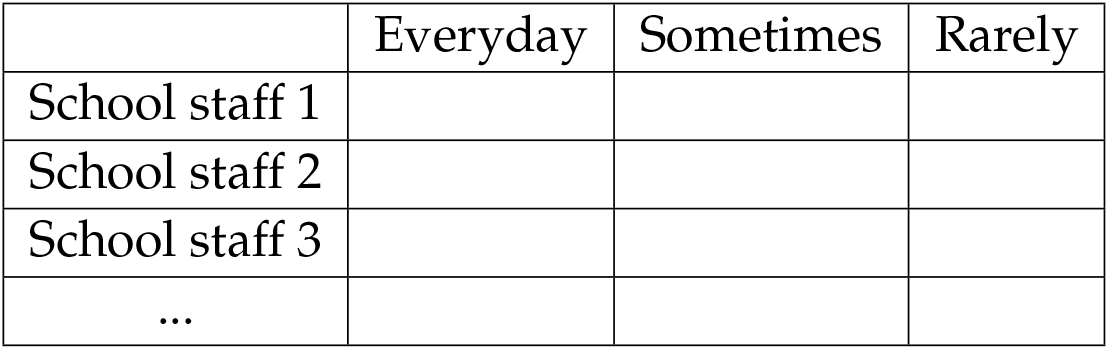

With whom from your colleagues do you spend time outside of school (e.g. on the way to work, free time)?

*In case you do not meet the person in a given row, do not check any value. Your name is also in the list, do not fill any value in that row*.

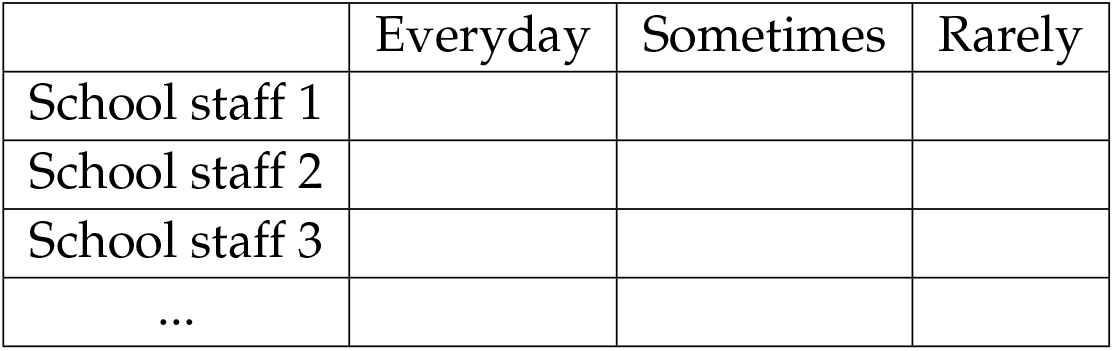

In which classes do you teach and how many lessons per week?

*In case you do not teach in a given class, do not fill any value*.

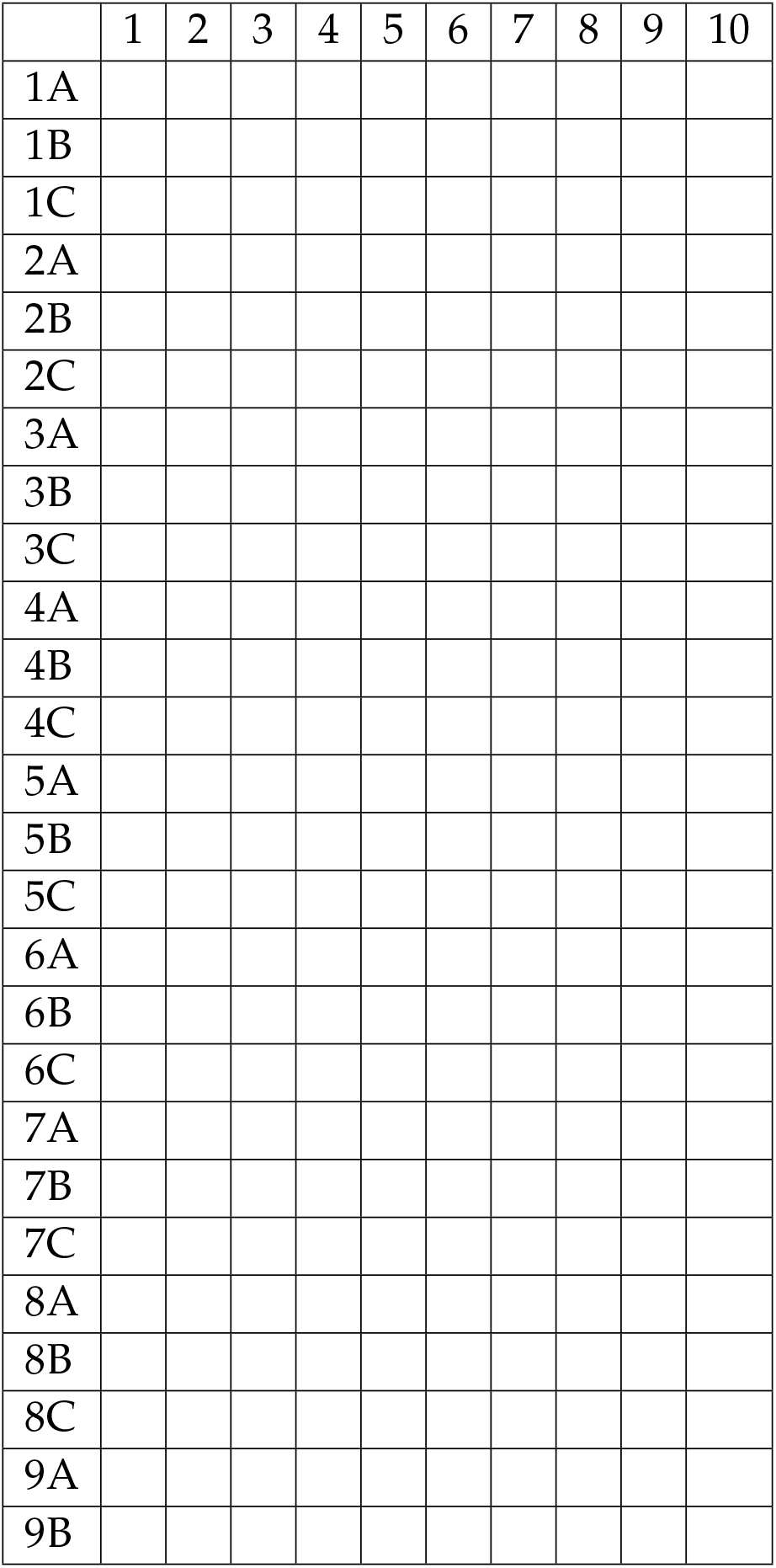

In which classes do you work as an educational assistant?

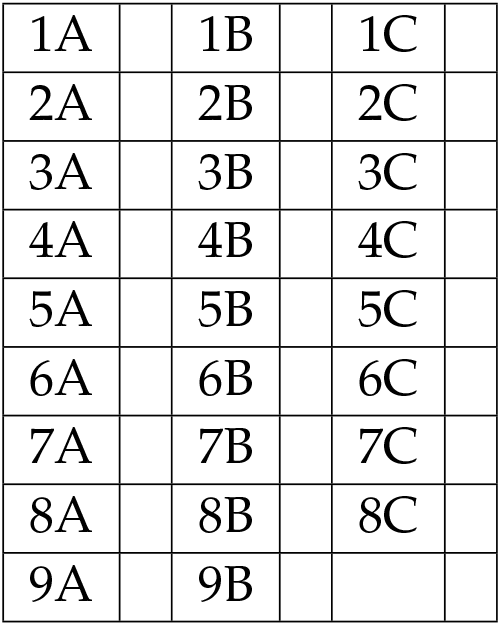

Pupils from which classes do you usually meet in your position as the after school club assistant?

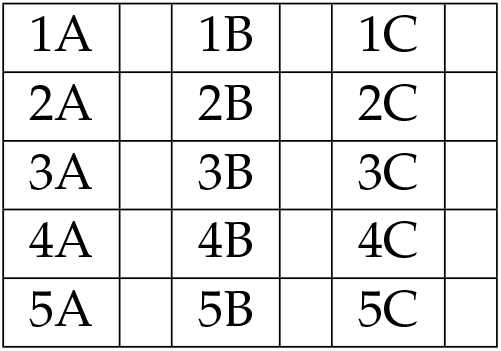

|  |  |  |  |  |
| --- | --- | --- | --- | --- |
| 1A |  | 1B |  | 1C |
| 2A |  | 2B |  | 2C |
| 3A |  | 3B |  | 3C |
| 4A |  | 4B |  | 4C |
| 5A |  | 5B |  | 5C |

